# HIV-phyloTSI: Subtype-independent estimation of time since HIV-1 infection for cross-sectional measures of population incidence using deep sequence data

**DOI:** 10.1101/2022.05.15.22275117

**Authors:** Tanya Golubchik, Lucie Abeler-Dörner, Matthew Hall, Chris Wymant, David Bonsall, George Macintyre-Cockett, Laura Thomson, Jared M. Baeten, Connie L Celum, Ronald M. Galiwango, Barry Kosloff, Mohammed Limbada, Andrew Mujugira, Nelly R Mugo, Astrid Gall, François Blanquart, Margreet Bakker, Daniela Bezemer, Swee Hoe Ong, Jan Albert, Norbert Bannert, Jacques Fellay, Barbara Gunsenheimer-Bartmeyer, Huldrych F. Günthard, Pia Kivelä, Roger D. Kouyos, Laurence Meyer, Kholoud Porter, Ard van Sighem, Mark van der Valk, Ben Berkhout, Paul Kellam, Marion Cornelissen, Peter Reiss, Helen Ayles, David N. Burns, Sarah Fidler, Mary Kate Grabowski, Richard Hayes, Joshua T Herbeck, Joseph Kagaayi, Pontiano Kaleebu, Jairam R Lingappa, Deogratius Ssemwanga, Susan H Eshleman, Myron S Cohen, Oliver Ratmann, Oliver Laeyendecker, Christophe Fraser, the HPTN 071 (PopART) Phylogenetics protocol team, the BEEHIVE collaboration and the PANGEA consortium

**Affiliations:** Big Data Institute, Nuffield Department of Medicine, University of Oxford, Oxford, UK; Wellcome Centre for Human Genetics, Nuffield Department of Medicine, University of Oxford, Oxford, UK; Department of Global Health, University of Washington, Seattle, WA, USA; Department of Medicine, University of Washington, Seattle, WA, USA; International Clinical Research Center, University of Washington, Seattle, WA, USA; Rakai Health Sciences Program, Entebbe, Uganda; London School of Hygiene and Tropical Medicine, London, UK; Zambart, University of Zambia, Lusaka, Zambia; Infectious Diseases Institute, Makerere University, Kampala, Uganda; Kenya Medical Research Institute (KEMRI), Nairobi, Kenya; European Molecular Biology Laboratory, European Bioinformatics Institute, Wellcome Genome Campus, Hinxton, Cambridge, UK; Centre for Interdisciplinary Research in Biology (CIRB), Collège de France, CNRS, INSERM, PSL Research University, Paris, France; IAME, UMR 1137, INSERM, Université de Paris, Paris, France; Department of Medical Microbiology and Infection Prevention, Molecular Diagnostic Unit, Academic Medical Center, Amsterdam, the Netherlands; Stichting HIV Monitoring, Amsterdam, the Netherlands; Wellcome Sanger Institute, Wellcome Genome Campus, Cambridge, UK; Department of Microbiology, Tumor and Cell Biology, Karolinska Institutet, Stockholm, Sweden; Department of Clinical Microbiology, Karolinska University Hospital, Stockholm, Sweden; Division for HIV and Other Retroviruses, Robert Koch Institute, Berlin, Germany; School of Life Sciences, Ecole Polytechnique Fédérale de Lausanne, Lausanne, Switzerland; Swiss Institute of Bioinformatics, Lausanne, Switzerland; Precision Medicine Unit, Lausanne University Hospital and University of Lausanne, Lausanne, Switzerland; Department of Infectious Disease Epidemiology, Robert Koch-Institute, Berlin, Germany; Division of Infectious Diseases and Hospital Epidemiology, University Hospital Zurich, Zurich, Switzerland; Institute of Medical Virology, University of Zurich, Zurich, Switzerland; Department of Infectious Diseases, Helsinki University Hospital, Helsinki, Finland; INSERM CESP U1018, Université Paris Saclay, APHP, Service de Santé Publique, Hôpital de Bicêtre, Le Kremlin-Bicêtre, France; Institute for Global Health, University College London, London, UK; Amsterdam UMC location Meibergdreef, Amsterdam, the Netherlands; Kymab Ltd, Cambridge, UK; Department of Infectious Diseases, Department of Medicine, Imperial College London, London, UK; Department of Global Health, Amsterdam University Medical Centers, University of Amsterdam and Amsterdam Institute for Global Health and Development, Amsterdam, the Netherlands; Division of AIDS/National Institute of Allergy and Infectious Diseases, National Institutes of Health, Rockville, MD, USA; Department of Infectious Disease Epidemiology, Imperial College, London, UK; Department of Epidemiology, Johns Hopkins Bloomberg School of Public Health, Baltimore, MD, USA; Department of Pathology, Johns Hopkins University School of Medicine, Baltimore, MD, USA; Institute for Disease Modeling, Bellevue, Washington, USA; Medical Research Council (MRC)/Uganda Virus Research Institute (UVRI), Uganda; London School of Hygiene and Tropical Medicine (LSHTM) Uganda Research Unit; Uganda Virus Research Institute, Entebbe, Uganda; Department of Pediatrics, University of Washington, Seattle, Washington, USA; Department of Medicine, University of North Carolina at Chapel Hill, Chapel Hill, NC, USA; Department of Mathematics, Imperial College, London, UK; Division of Intramural Research, National Institute of Allergy and Infectious Disease, National Institutes of Medicine, Baltimore, MD, USA

## Abstract

Estimating the time since HIV infection (TSI) at population level is essential for tracking changes in the global HIV epidemic. Most methods for determining duration of infection classify samples into recent and non-recent and are unable to give more granular TSI estimates. These binary classifications have a limited recency time window of several months, therefore requiring large sample sizes, and cannot assess the cumulative impact of an intervention. We developed a Random Forest Regression model, HIV-phyloTSI, that combines measures of within-host diversity and divergence to generate TSI estimates from viral deep-sequencing data, with no need for additional variables. HIV-phyloTSI provides a continuous measure of TSI up to 9 years, with a mean absolute error of less than 12 months overall and less than 5 months for infections with a TSI of up to a year. It performed equally well for all major HIV subtypes based on data from African and European cohorts. We demonstrate how HIV-phyloTSI can be used for incidence estimates on a population level.

## Introduction

Accurate estimates of HIV incidence are critical for surveillance of HIV epidemics and to determine the effectiveness of prevention efforts (Mastro 2013). HIV incidence, the rate at which new infections arise from the susceptible part of the population, tracks the leading edge of the epidemic, measuring transmission at a given moment in time. The traditional longitudinal cohort approaches for measuring incidence are costly, and incidence derived from a cohort with many HIV prevention interventions is likely to give biased estimates (Incidence Assay Critical Path Working Group 2011; Brookmeyer 1997; Busch et al. 2010).

A cross-sectional study design would be preferable for measuring incidence, and recent methodological advances have brought this possibility closer. Cross-sectional HIV incidence estimation is currently based on algorithms that use observable changes in the biomarker profile of individuals to class infections as recent or non-recent (Busch et al. 2010). Identification of individuals with recent HIV infection allows one to estimate incidence by counting the number of such individuals within the population/cohort and adjusting for the time period associated with that recent state. Using the quality of the antibody response to HIV infection as a host biomarker, these methods have been used to estimate HIV incidence at a country level (Voetsch et al. 2021), within specific risk groups (Teixeira et al. 2021), as the primary outcome of intervention trials, and have been considered for counterfactual incidence estimation to determine the effectiveness of Pre-Exposure Prophylaxis (PrEP).

An alternative to host response biomarkers is measurement of the genetic diversity of HIV sequences found in individual infections. It has long been recognised that in the absence of treatment, HIV viral genetic diversity increases with duration of infection in a single host (Shankarappa et al. 1999; Zanini et al. 2015), and several studies have used measures of HIV genetic diversity to estimate HIV incidence. These studies use several sequence-based and non-sequence-based parameters: nucleotide ambiguity (Kouyos et al. 2011; Ragonnet-Cronin et al. 2012), Hamming distance - Q10 (Park et al. 2011), generalized entropy (Wu et al. 2015), pairwise distances (Moyo et al. 2016), time to most recent common ancestor (MRCA) (Moyo et al. 2017) and high-resolution melting diversity assays (Cousins et al. 2014). However, no single approach has demonstrated sufficient precision for HIV incidence estimation.

In recent years, next-generation sequencing (NGS) data has shown promise for identifying recent infections from genetic sequence data alone. Puller et al showed that a new NGS-based measure (3rd base ambiguity in the *pol* gene) can be used to estimate time since HIV-1 infection (TSI) many years after the infection, in contrast to most alternative biomarkers. The study was based on data from 42 patients in (Puller, Neher, and Albert 2017) and is yet to be validated in a larger population. TSI has also been estimated using average pairwise diversity (Carlisle et al. 2019), and a combination of pairwise sequence diversity and divergence in sub-sequences within *pol* and *env* in 30 individuals (Zhou et al. 2021). These studies also show that sequence methods have the potential to estimate TSI for the entire duration of infection, rather than classifying individuals as ‘recent’ or ‘non-recent’. A seminal study from 1999 demonstrated that “DNA Distance”, or longest root to tip distance (LRTT), increased almost linearly with time since infection (TSI) (Shankarappa et al. 1999; Zanini et al. 2015). However, LRTT relies on multiple sequences from a single time point, which historically has been costly to compute and therefore seen as impractical for population studies. This is changing with the widespread adoption of next-generation sequencing.

In this study, we present a new method for estimating TSI of all major HIV-1 subtypes from deep sequence data, using both LRTT and diversity: HIV-phyloTSI. HIV-phyloTSI employs a machine-learning algorithm, using data from four different cohorts representing 480 individuals with known approximate TSI as a training dataset. Performance was evaluated in two ways: first, using a simulated dataset, and second, using a dataset from the HPTN-071 (PopART) Phylogenetics Ancillary study (Hall et al. 2021) (“HPTN 071-02 Study Protocol” 2017) The output of HIV-phyloTSI are continuous estimates of TSI, rather than a binary classification of samples as recent and non-recent, and the method is sufficiently accurate to estimate HIV incidence in a cross-sectional cohort without use of additional demographic variables.

## Results

### Participants and datasets

HIV genome sequences were made available by the PANGEA and BEEHIVE consortia following consortium-specific guidelines for data access. PANGEA (Abeler-Dörner et al. 2019) is a network of collaborators from Africa, Europe and the United States (US) who have generated a large number of HIV NGS sequences from Eastern and Southern Africa. The samples used for this study were collected in Uganda, Kenya and South Africa by MRC Uganda/UVRI (Asiki et al. 2013; Kasamba et al. 2019; Asiki et al. 2011) (MRC), the University of Washington International Clinical Research Centre programs (Celum et al. 2010; Baeten et al. 2012; Lingappa et al. 2011) (UWP) and the Rakai Community Cohort Study (Wawer et al. 1999; Chang et al. 2016) (RAK). BEEHIVE (Blanquart et al. 2017) (BEE) is a network of HIV cohorts that collected HIV samples from HIV seroconverters across Western Europe (Belgium, Finland, France, Germany, the Netherlands, Sweden, Switzerland, United Kingdom) and in Uganda. Participants in all studies were treatment-naive and viraemic at the time of sampling. Participants in all studies gave written consent and ethics approvals were granted to the institutions that generated the data. Participant characteristics for the different datasets are listed in Table 1.

**Table 1:**
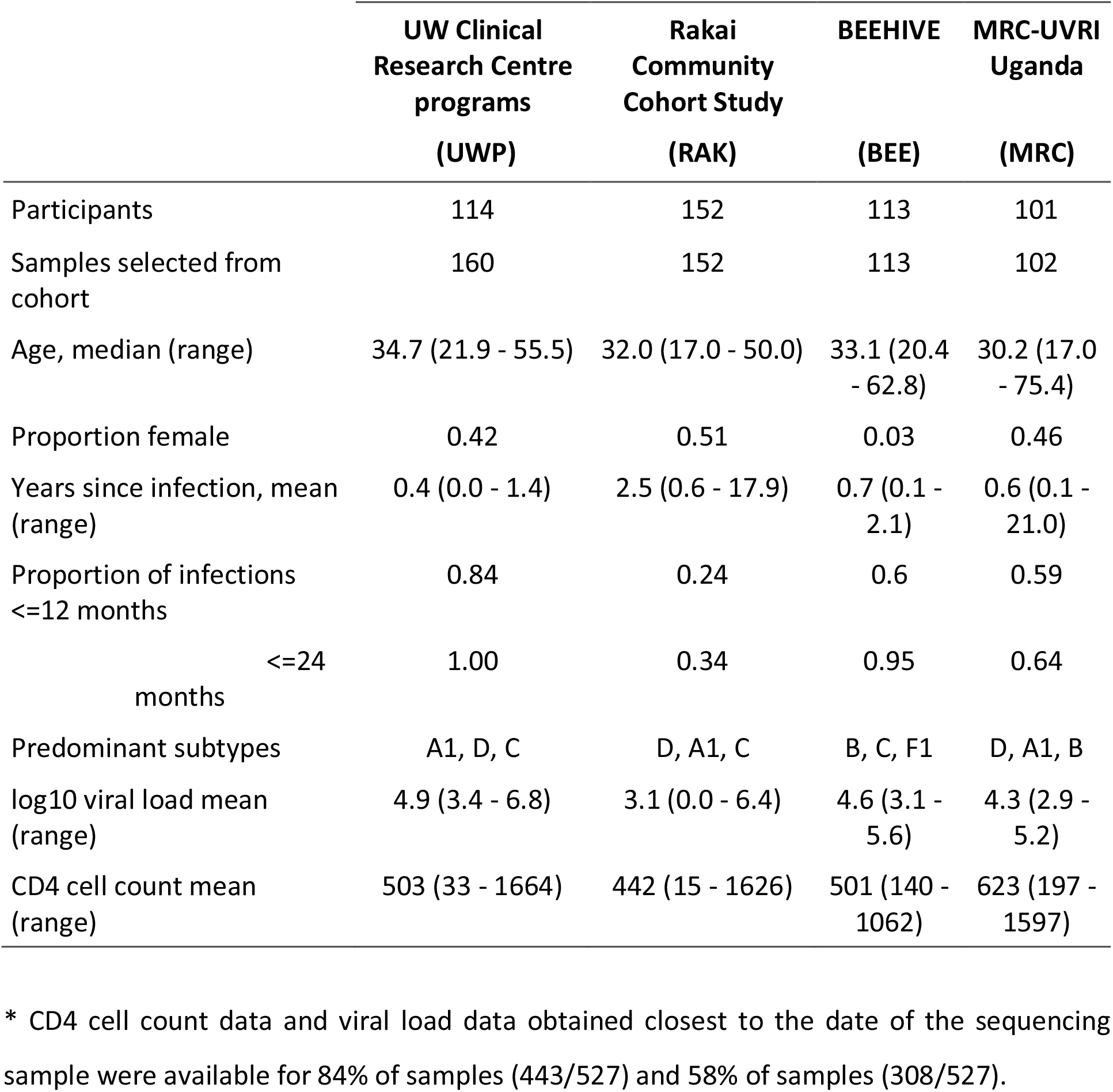
Participant characteristics in training dataset.

Datasets were chosen to include participants with known seroconversion intervals (UWP and BEE) and participants with recent as well as participants with non-recent infections (RAK and MRC) (Figure 1). No other pre-selection was made, but as only sequenced genomes were included, this of necessity implies detectable viral load (above 10^2^ copies per ml). Cohorts from different geographical locations were included to ensure a representation of subtypes A, B, C, and D. In total, the training dataset consisted of 527 sequences from 480 participants (Table 1).

**Figure 1.**
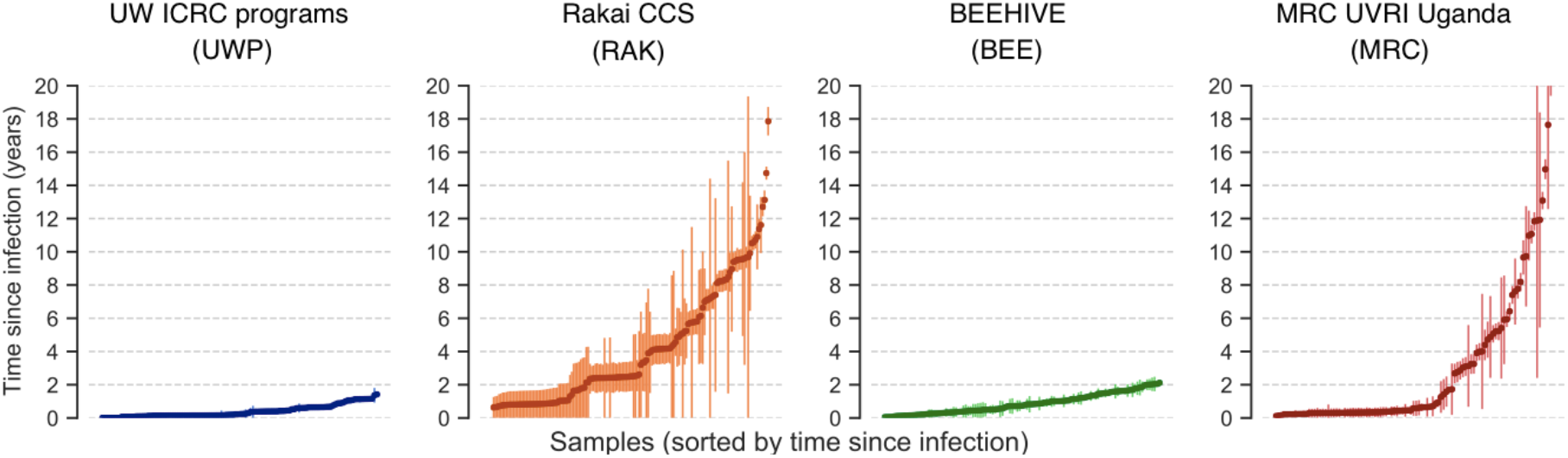
Distribution of time since infection (TSI) and seroconversion intervals in the training datasets. Vertical lines show duration of the seroconversion interval, from three weeks prior to the last HIV-negative test to the date of the date of the first HIV-positive test. TSI was defined as the time in years between the sampling date and the midpoint of the seroconversion interval (circles).

### HIV diversity and divergence increase with the duration of infection

Minor allele frequency (MAF) has been shown to increase over the course of an HIV infection and has previously been used to identify recent infections (Carlisle et al. 2019; Puller, Neher, and Albert 2017; Ragonnet-Cronin et al. 2012, 2021; Kouyos et al. 2011). In this study, we split MAF into two variables, designated MAF12c for the first two codon positions and MAF3c for the third codon position, to allow for their different evolutionary rates. Both MAF12c and MAF3c increased with TSI (Figure 2), at rates that varied by HIV gene. The most rapid increase was observed for MAF3c in the *gp120* and *gp41* regions, this is consistent with the high substitution rate in the *env* gene relative to the more constrained *pol* and *gag* genes. MAF3c was informative across the genome, particularly in *gag, pol* and *gp120* (Figure S1), while MAF12c remained highly constrained in *gag* and *pol*, but was markedly more informative in *gp120* (Figure S1). We hypothesized that estimates could be improved by using more complex statistics derived from phylogenetic trees. We therefore constructed intrahost diversity trees in sliding windows along the genome using *phyloscanner* (Wymant, Hall, et al. 2018) and tested if pairwise patristic distance, overall root-to-tip length or longest root-to-tip distance (LRTT) were predictors of TSI (Figure S2). LRTT performed best, and was included alongside MAF12c and MAF3c as a predictor in this study. LRTT was particularly informative in *gp120* and for non-recent infections (Figure 2). The performance of all three predictors was not influenced by the sequencing method used (amplicon-based or probe-based) (Figure S3).

**Figure 2.**
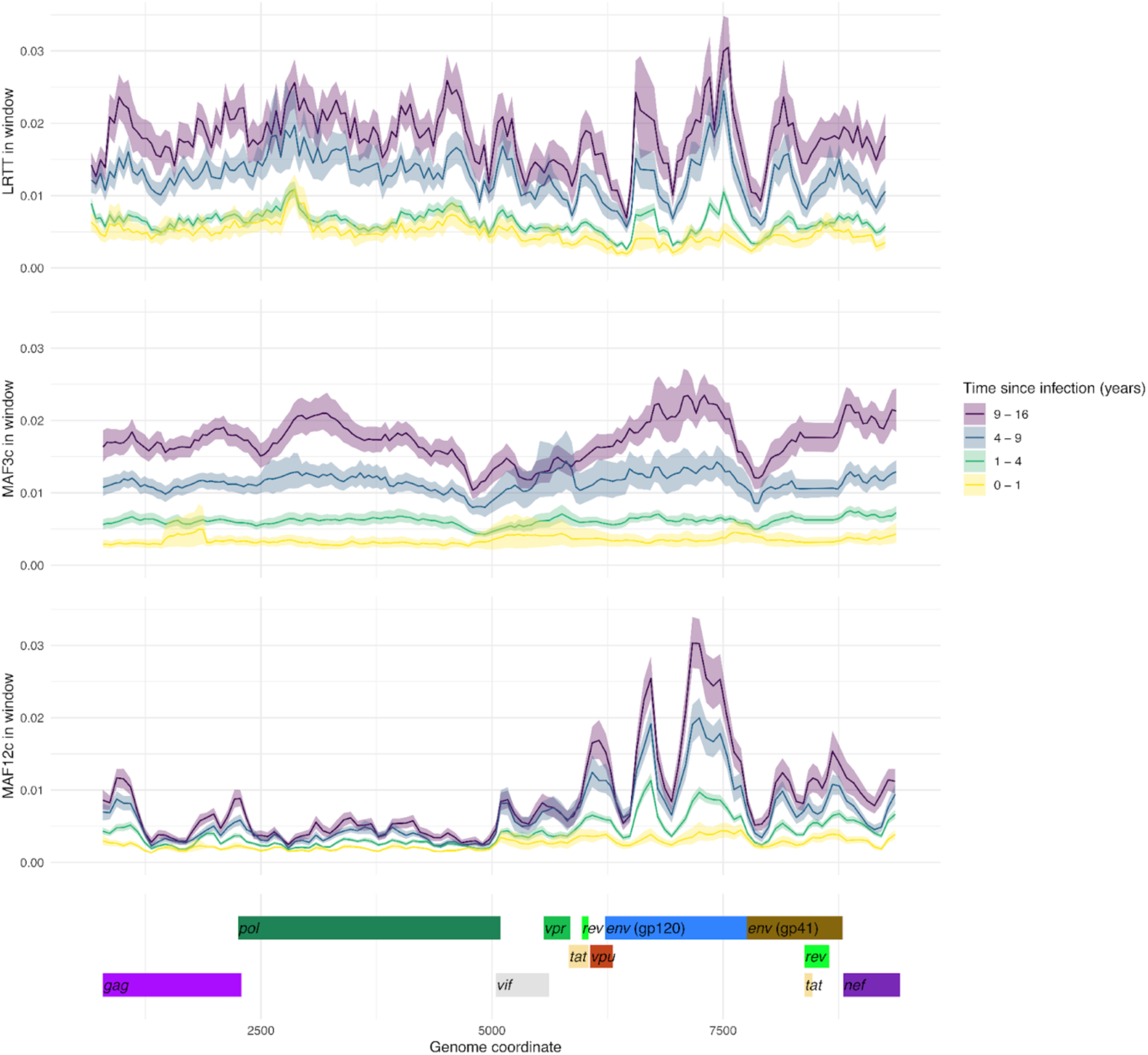
Genetic divergence (LRTT) and diversity (MAF3c, MAF12c) values along the HIV genome in samples grouped by time since infection. Lines show mean root-to-tip distance of the largest subgraph (LRTT), and mean minor allele frequencies at the third codon position (MAF3c) and first and second positions (MAF12c). MAF3c and MAF12c measures shown as mean per 250 bp window. The shaded area indicates the width of the 95% bootstrap confidence interval around the mean.

To determine whether some genomic locations were particularly informative, we assessed the correlation of MAF12c, MAF3c, and LRTT values across the HIV genome with the known TSI (Figure S4). Informativeness was more variable for the MAF predictors than for LRTT. MAF3c was most informative in the 3’ region of *gag*, the 5’ region of *pol*, and the 5’ region of *gp120*; MAF12c was most informative in the 5’ region of *gp120*; and LRTT was informative in all of *gag, pol* and *gp120*. The most informative windows, defined as having r2 over 0.3 for any predictor, were included as one of the feature combinations during model selection (LRTT_MAF3c_topwin). MAF3c alone was highly informative, but performance was improved by including other parameters.

In conclusion, LRTT adds a measure of TSI that is complementary to MAF. Both predictors perform best in the *gp120* region of the *env* gene, however, all regions of the genome are informative. A model should ideally aggregate information across all genomic regions to become more powerful.

### CD4 cell counts and age are informative but are not essential for estimating TSI

Limited clinical data were available for the sequences in the training dataset. CD4 cell counts and viral load measurements obtained close to the date of the sequencing sample were available for 84% (443/527) and 58% (308/527) of the samples, respectively. Mean CD4 cell count and mean log10 viral load were significantly different between recent and non-recent infections (CD4 count, 543 v 451 cells/mm3; log10 viral load, 4.9 v 4.3; *p*< 0.001, Welch’s t-test, with recency cut-off of 12 months) (Figure 3A,B), but with high variance and substantial overlap between the distributions. In datasets where CD4 cell count data are available from the sample collection date, recency estimates may be further improved by incorporating this variable. In the present dataset, gains were marginal (<1 percentage point in r^2^). Age was informative in the two population cohorts (RAK and MRC), but not in the two cohorts enriched for seroconverters (UWP and BEE) (Figure 3C). We opted against including age as a variable in the present study so as to keep the model as generally applicable as possible.

**Figure 3.**
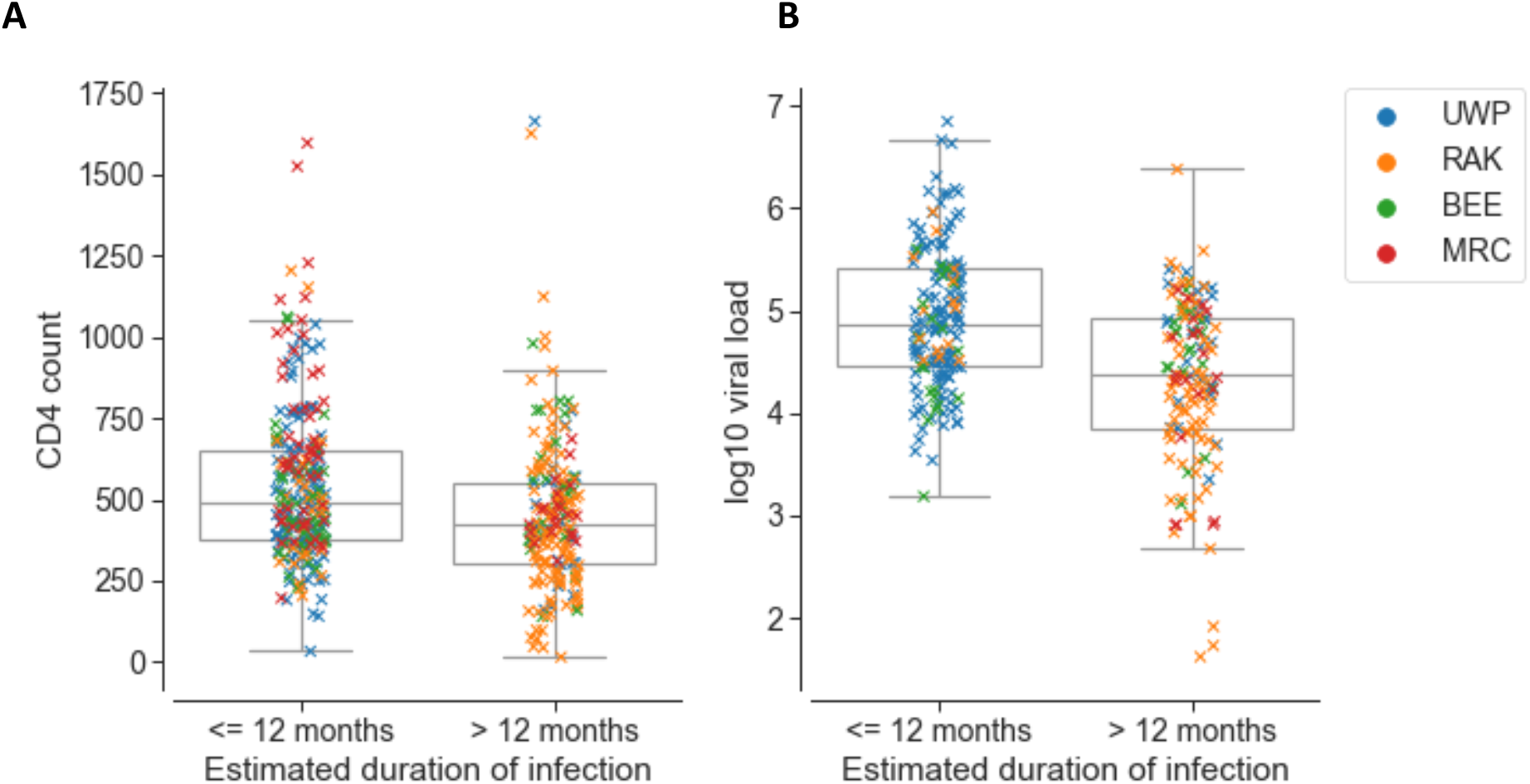
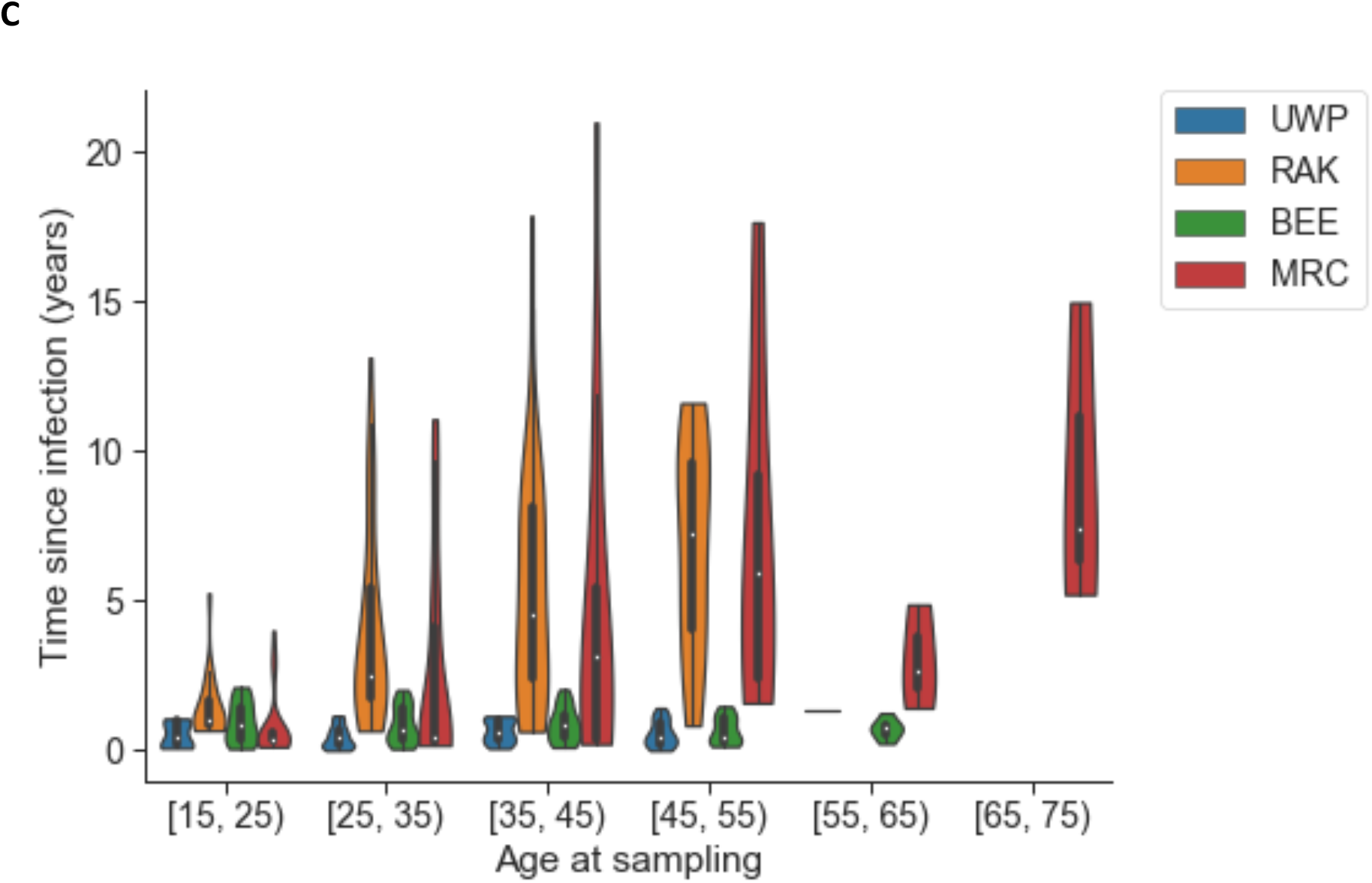
CD4 cell count, viral load and age can be informative. A. Most recent CD4 cell count obtained closest to sequencing sampling date. B. Viral load (log10 copies per mL) obtained closest to sequencing sampling date. C. Distribution of age at sampling in different cohorts in the training data set.

The estimates were not influenced by the presence of drug resistance mutations. Given the dates of sampling, none of the participants was infected while using PreP.

### Models based on diversity and divergence yield good accuracy and false recency rates

The chosen predictor variables - MAF12c, MAF3c and LRTT - were calculated for each window of 250 base pairs and then averaged separately across *gag, pol, env gp120* and *env gp41*, as well as across the whole genome. In five samples, missing data meant that gene-averaged values could not be derived for one or more genes; these were filled by imputation. To select a regression method, we tested performance of three commonly used methods (ordinary least squares (OLS), gradient boosted regression, and random forest regression), using a subset of feature combinations to estimate the square-root transformed TSI. The square-root transformation helped the model accommodate greater uncertainty around the actual TSI values (wider seroconversion intervals) in non-recent infections (Figure 1). Cross-validated model scores (mean r^2^ values) were computed using 10-fold cross-validation, in each iteration splitting the data randomly into 75% training and 25% test datasets and scoring model performance on the test data (Table S1).

OLS performed adequately for the simplest models, where a single feature was used in addition to the indicator variable for sequencing method, but was outperformed by gradient boosted and random forest regression for larger feature sets. The cross-validated r^2^ scores were similar for gradient boosted and random forest regression, with random forest slightly outperforming gradient boosted regression on all feature combinations assessed (Table S1). Scatterplots of known versus estimated square root-transformed TSI for each of the feature combinations are shown in Figure S5. Random forest regression was selected for subsequent analyses.

To assess model performance with different combinations of features, we compared several metrics using 20-fold cross validation: mean r^2^ on test data (strength of correlation with known TSI) (Figure 4A), accuracy of predicting infections as having TSI below either 12 or 18 months (Figure 4B), mean absolute error (MAE) (Figure S6A) and the false recency rate (FRR) (Figure S6B). Since our model predicted continuous TSI, we were able to obtain binary estimates (recent/non-recent) at any arbitrary recency cut-off after running the model. We compared this continuous approach to a ‘classification’ approach that uses a binary outcome throughout, based on a pre-decided threshold of recent versus non-recent infections. Specifically, we used the same feature sets to fit random forest classifier models, with recency defined as true TSI below either 12 or 18 months. For a TSI threshold of 12 months, regression with discretisation at 12 months outperformed classification for all feature sets (Figure S7). For a TSI threshold of 18 months, classification showed an improvement for the most complex feature sets that included individual genomic windows (Figure S7B), but without a corresponding improvement in overall accuracy (Figure S7A). We used the regression approach in subsequent analyses.

**Figure 4.**
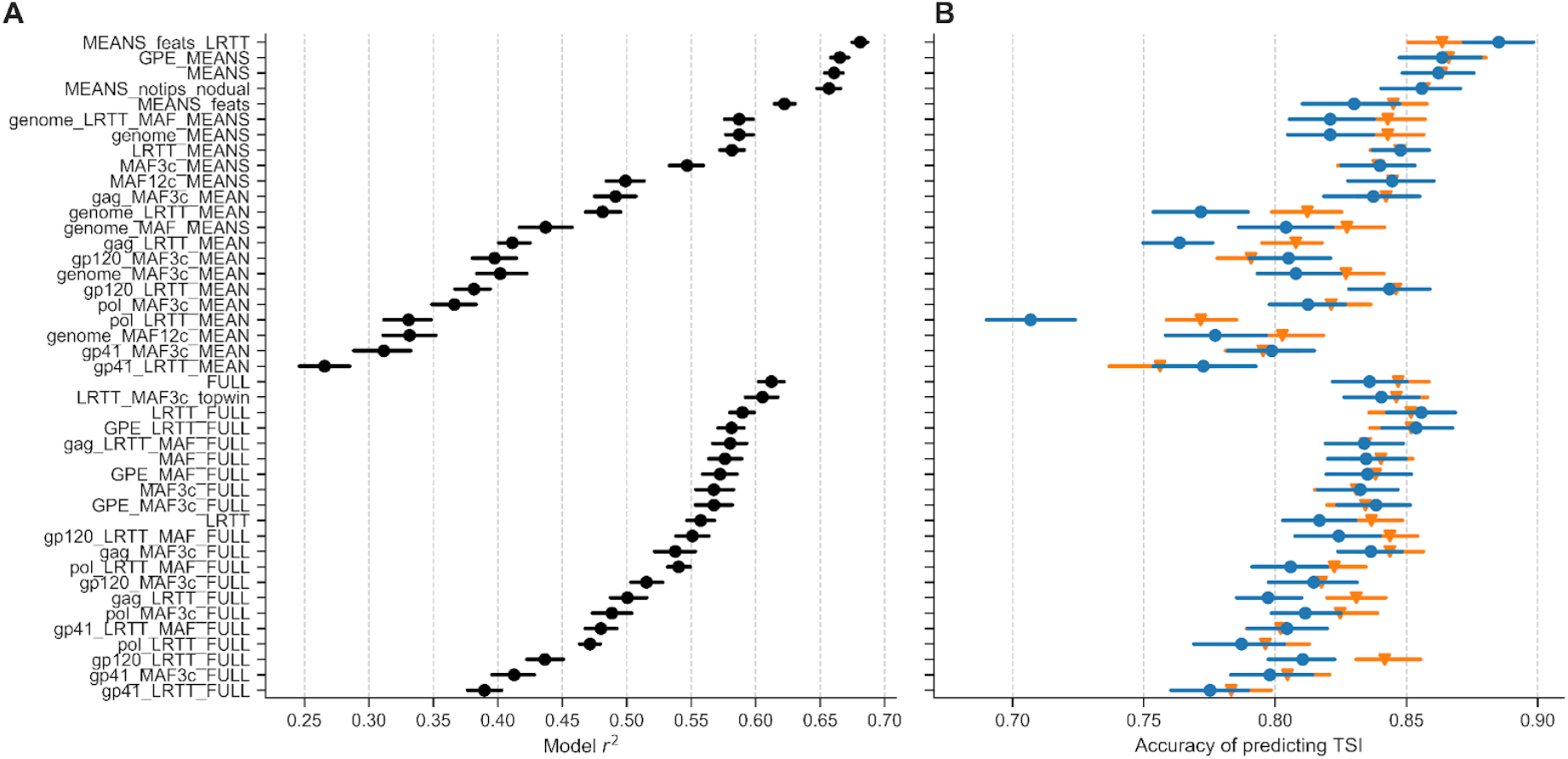
Model r^2^ and accuracy of identifying recent infections. Markers show mean and lines show 95% bootstrap CI over 20-fold cross-validation. A. Model r^2^ score on test data. B. Accuracy, defined as the proportion of samples having TSI correctly estimated as being below or above a cut-off of either 12 months (circles, blue) or 18 months (triangles, orange).

Feature sets either used information from individual genomic windows (‘FULL’ in feature set name), or aggregated information from all windows covering a given gene or the whole genome (‘MEAN’ in feature set name). Feature aggregation resulted in a gain of power: models built with aggregated feature sets had higher r^2^ scores and accuracy than those built with individual genomic windows (Figure 4). Accuracy varied depending on how genomic features were aggregated (Figure 4). This was mostly due to missing data. For the same feature combinations, the best-performing aggregate-feature model MEANS_feats_LRTT, which comprised ten aggregated features across *gag, pol, gp120* and *gp41* as listed in Table S2, had a cross-validated r^2^ score of 0.68 and accuracy of 0.89, while the best-performing individual-windows model FULL had an r^2^ score of 0.61 and accuracy of 0.84. The overall best performing models were generated by the feature set “MEANS_feats_LRTT”, with the greatest proportion of variance explained by MAF and LRTT in the *gag* region, followed by LRTT in *gp120* and *pol* (Figure S8). The indicator variable for the type of sequencing, *is_mrc*, carried relatively little importance, suggesting that the results remain robust to the type of sequence data (amplicon-based or capture-based).

In conclusion, random forest regression was chosen over OLS and gradient boosted regression, and a continuous regression over a binary classification. Models with aggregated feature sets performed better than those using individual genomic windows. The best performing aggregated feature set, MEANS_feats_LRTT, was used to fit the final predictive model.

### HIV-phyloTSI is suitable for population-level predictions

We evaluated the final modzel on the original dataset using a leave-one-out strategy on the original dataset (Figure 5), on a simulated population (Figure 6) and on an independent dataset (Figure 7).

**Figure 5:**
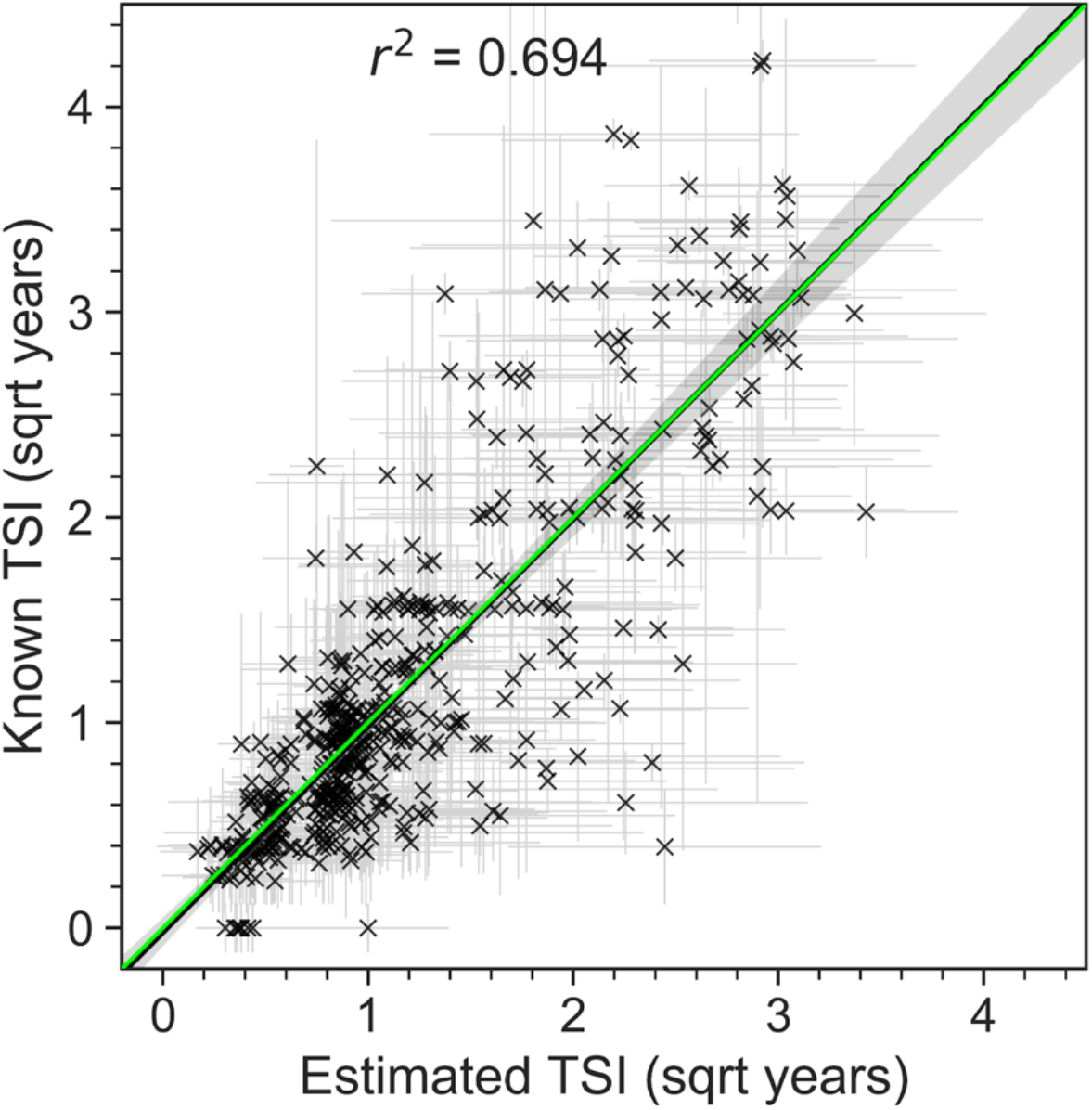
HIV-phyloTSI is more accurate on population level. A. TSI estimates compared with known midpoint TSIs, with point estimates indicated as crosses and seroconversion intervals as lines. Regression line shown in black with confidence interval as shaded grey area. The green line (overlapping with the black line) indicates equality.

**Figure 6:**
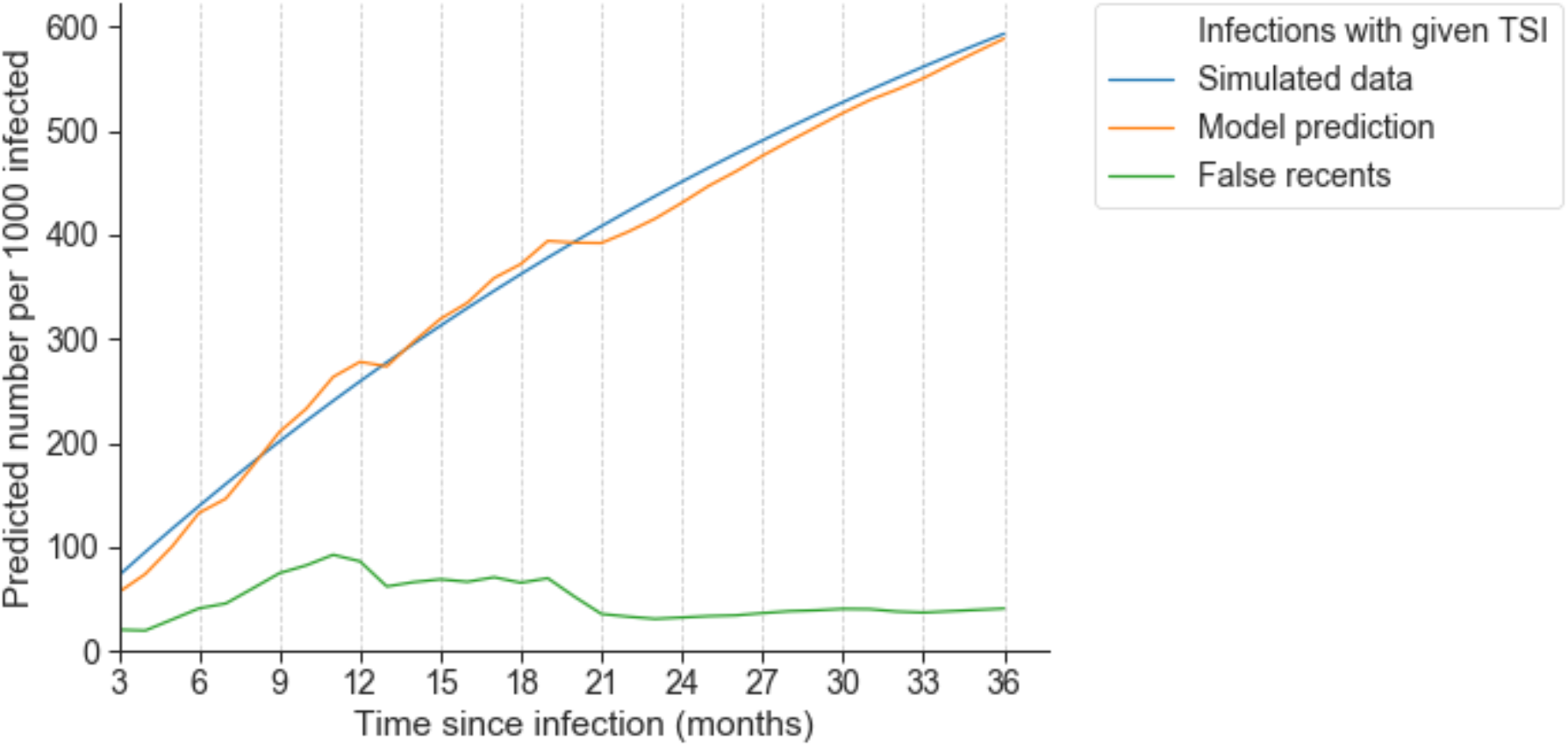
HIV-phyloTSI can predict recency at different cut-offs in a simulated population. Number of recent infections in a simulated population of 1000 individuals where average time to treatment is 3 years, with the recency cut-off varied between 3 and 26 months. The blue line shows the true number; the orange line shows the number estimated by HIV-phyloTSI; the green line shows the subset of estimated recent infections that were falsely estimated to be recent (i.e., were in truth non-recent).

**Figure 7:**
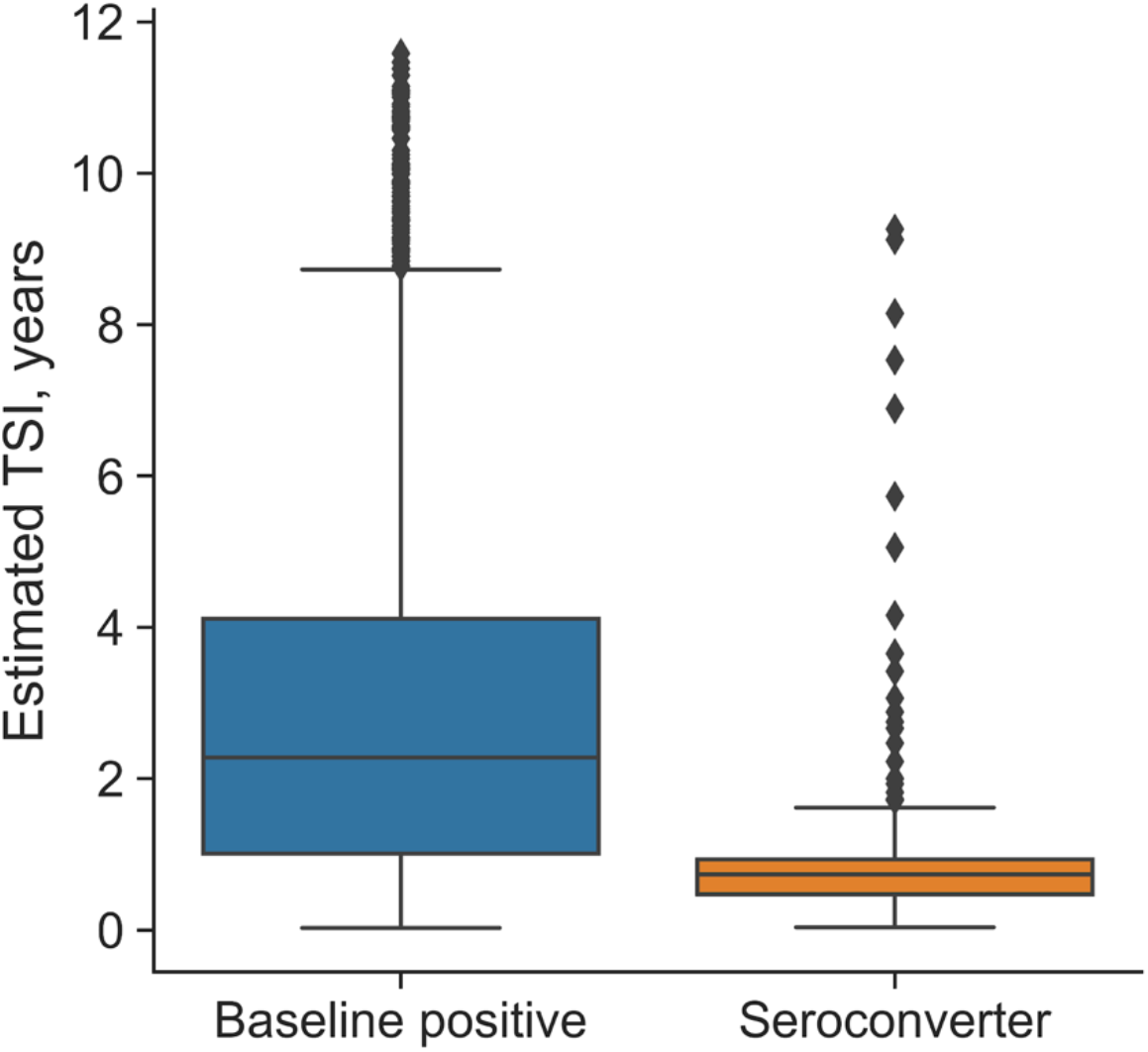
HIV-phyloTSI is suitable for population-level predictions. Distribution of TSI estimates for members of the PopART Population Cohort (PC), by HIV status. ‘Baseline positive’ indicates individuals positive for HIV at enrolment (1041 samples); ‘seroconverter’ indicates individuals who acquired HIV during the study period (204 samples). Boxes show extent of first quartile from the median (line); whiskers extend to 1.5*IQR.

Estimates of TSI were made for all 527 samples in a leave-one-out strategy, iterating over the full sample set, building a random forest model on the other 526 samples with the previously selected parameters, and obtaining a point estimate together with the full distribution of estimates from each decision tree in the forest. A separate random forest regression model was trained on the mean absolute errors of predictions from the base model, to generate prediction intervals around the estimates (Figure 5).

Figure 5 shows the overall fit for the best performing model with feature set MEANS_feats_LRTT, comparing known seroconversion intervals against TSI estimates and prediction intervals from the model. The overall regression line is close to the line of equality, indicating that over the majority of TSIs, recency and non-recency are misclassified to the same extent, despite a tendency to slightly overestimate TSIs below 1 year, and underestimate the TSIs greater than 9 years (Figure S9). This implies that although precision is insufficient for an individualised clinical assay, population-level incidence estimates are likely to be accurate, at least in datasets with similar distribution of recency.

The suitability of HIV-phyloTSI for population analysis was tested by applying the model to a simulated population (Figure 6, Table S2). One thousand people were drawn from a population with an average interval of three years from infection to treatment. The duration of infection of individuals in this population with TSI *w* was modelled as 1 - e^-0.3w^. The predicted fraction of infections with estimated TSI up to *w* in this population, and the proportion of these that were incorrectly classified, were calculated using the false and true recency rates for the model at values of *w* between 3 and 36 months. Plotted is the number of recent infections out of the 1000 people depending on where the cut-off for recency is set. The model predictions (orange line) closely track the numbers expected from the simulated data (blue line)

Next, we applied the model to an independent dataset, namely the HPTN 071-02 (PopART) Phylogenetics ancillary study (“HPTN 071-02 Study Protocol” 2017) (Figure 7). The study generated HIV1 sequences from HPTN 071 trial participants in Zambia (Hayes et al. 2019). We sequenced samples from 204 participants who were HIV negative at enrolment and seroconverted both during the trial and less than one year after their last negative test, shown in orange, and 1041 participants who were positive at enrolment, shown in blue. Most, but not all of these participants were likely infected for more than a year before enrolment. HIV-PhyloTSI results reveal a highly significant difference between the distribution of TSI for samples in the two groups (p<<0.001, Welch’s t-test). The median estimated TSI for baseline positives was 2.28 years (interquartile range 1.01-4.11) and for seroconverters 0.74 years (IQR 0.47-0.94). Of the 204 seroconverters with known seroconversion intervals, 7 (3.4%) were incorrectly predicted based on a lower TSI limit of over 12 months.

In conclusion, HIV-phyloTSI is able to predict recency with sufficient accuracy in population-level analyses.

### HIV-phyloTSI is able to predict recency of infection for all subtypes

Recency assays based on MMAs have shown variable performance for different HIV-1 subtypes (Longosz et al. 2014). We therefore tested whether the model performance showed bias in predictions for any subtype. Our dataset included at least 100 samples for each of subtypes A-D (A1, 138; B, 101; C, 232; D, 147) as well as small numbers of other subtypes and circulating recombinant forms, which we grouped for this analysis (“Other”, 113). Adjustment for TSI was required since model error increases with TSI regardless of subtype (Figure 8A and S9), and the over-representation of subtypes A1 and D among non-recent samples in our dataset would otherwise inflate the error range for these subtypes. After adjusting for TSI, there was no difference in bias for any of the subtypes (Figure 8B and Table S3, p>0.05 for every pairwise comparisons, Tukey’s range test), indicating that model performance was independent of subtype.

**Figure 8:**
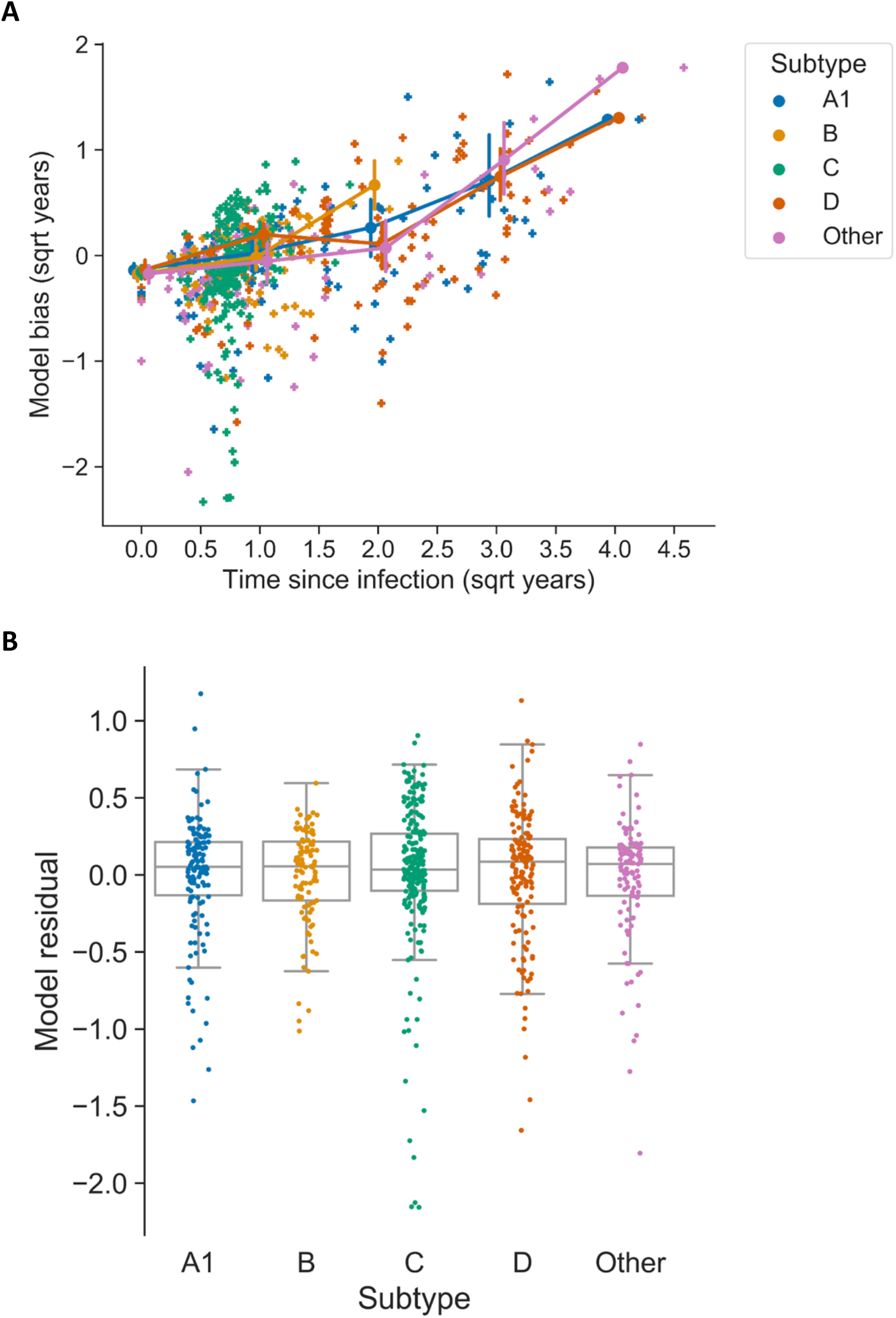
HIV-phyloTSI is unbiased with respect to subtype. A. Model bias (difference between real and estimated TSI value) by square-root transformed time since infection, coloured by subtype. Circles indicate mean for groups of 0-1, 2-4, 5-9 and 10-16 years since infection, and vertical bars indicate the 95% bootstrap confidence interval in each group, for all subtypes. Lines connect group means to aid visualisation. B. Residuals from the linear model used to adjust for TSI of samples (Bias ∼ TSI), shown by subtype, for subtypes with at least 5 samples in the dataset. Boxes extend to first quartile from the median (line) and whiskers to 1.5*IQR. Model bias increases with TSI for all subtypes, but is not significantly different between subtypes (p>0.05 for pairwise comparisons, Tukey range test).

## Discussion

In this study, we present a method that uses viral genetic diversity (MAF) and divergence (LRTT) to estimate a continuous measure of time since infection (TSI) from a single HIV-1 NGS sequencing sample per patient. Any value of TSI can be chosen as a threshold to give a binary recency assay, with an accuracy of up to 89%. Additional metadata can be used if available for a specific setting, but is not required to make predictions. The method works equally well on HIV-1 subtypes A-D and is accurate enough for population-level estimates of incidence.

HIV-phyloTSI is a generic method derived using samples from a range of populations in Western Europe and Eastern and Southern Africa. It was developed for population-level analyses performed in the PANGEA-HIV consortium (Abeler-Dörner et al. 2019). When used as a standalone tool, it enables the inference of epidemiological information without requiring extra participant data. The model can be further extended to incorporate moderately informative variables such as CD4 cell count and viral load. For sequences obtained with veSeq-HIV, viral load estimates can be obtained directly from counts of uniquely mapped sequencing reads, offering an additional sequence-derived parameter without the need for additional testing (Bonsall et al. 2020; Fogel et al. 2020). Although CD4 cell count and viral load provided no substantial gain in accuracy with our training dataset, these could offer higher gains for studies where these data are available from the same date as the sequencing samples. Lundgren et al recently explored the use of biomarkers to augment phylogenetic TSI estimates (Lundgren et al. 2021).

There was a slight (<1 percentage point in r^2^) improvement when the model was extended with the addition of age at sampling, but there are good reasons to avoid including demographic markers in a generic model, as this may bias the model towards the type of population used for training. In our case, the training data contained two large seroconverter cohorts (BEE, UWP) and the association with age was only evident in the population cohorts (RAK, MRC). Likewise, we did not include subtype in the model, to avoid spurious association with subtypes preferentially represented in the seroconverter cohorts and to maintain the generalisability of the model.

HIV-phyloTSI performs best when used for population-level predictions, where its accuracy is sufficiently high that the total number of infections identified with any given TSI corresponds closely with the expected number of infections at that TSI. The close correspondence between expected and observed population-wide estimates of recency is particularly useful, as it allows the identification of recent infections not only in growing epidemics in which recent infections are common, but also in declining epidemics where recent infections are substantially outnumbered by non-recent infections. Although HIV-phyloTSI can be used to give an estimate of the duration of infection on an individual level, the uncertainty on individual level is likely to be too high for clinical applications. The level of intrahost diversity as a proxy for duration of infection however is informative on an individual level and might become relevant for clinical management and potential HIV cure once treatment goes beyond classical ART. The size of the viral reservoir is lower in recently infected individuals compared to those in the chronic stage of infection, and thus easier to eradicate the virus (Jain et al. 2013). The utility of knowing the duration of infection at an individual level is useful for HIV cure protocols (Richman et al. 2009), and staging of individuals with recent infection is currently being used in clinical HIV cure trials (Fidler et al. 2020).

The method has limitations. Any machine-learning model depends on how well classified the training data set is, and the large uncertainties in the seroconversion dates of non-recent samples in our training datasets necessarily limit the accuracy of the model. The training data was selected from different cohorts in sub-Saharan Africa, but validation is still needed to establish that the results are further generalisable. The method also requires sufficient virus from an infection to generate not only a consensus genome but also accurately represent intrahost diversity. Viral loads in this study (2-7 log10 copies per mL) are representative of the majority of viraemic individuals. However, in a real population in the era of universal testing and treatment, a substantial number of individuals will be virally suppressed, and a substantial proportion of these may be incident infections. At present, incidence surveys such as PHIA (Saito et al. 2017) assume that all virally suppressed infections are non-recent, which tends to underestimate incidence in settings where ART may be initiated early in infection. Ideally, recency estimates from viraemic individuals would be combined with multi-assay algorithms (MAA) that generate recency estimates of individuals with viral suppression, to yield more robust population estimates of incidence, as pioneered by Ragonnet-Cronin et al (Ragonnet-Cronin et al. 2021). We and others are currently working on methods to obtain full-length or near-full-length HIV-1 genomes from samples with a low viral load. Finally, the requirement for deep-sequence data carries a substantial laboratory and computational cost. However, sequencing is rapidly becoming more available and affordable in low-income countries, and the cost of computational power is declining.

We did not test HIV-phyloTSI on sequences obtained by proviral sequencing from individuals with natural or ART-induced viral suppression. The effects of ART on within-host HIV-1 diversification and divergence are not well studied, but it is to be expected that both will be profoundly reduced. A similar effect has been described for PreP in two small-scale studies, one in macaques and one in humans (Zheng et al. 2012; Ruone et al. 2016). Both studies find reduced diversity in the HIV viral population four and ten months after infection while taking PreP, respectively. More research is needed to adjust sequence-based methods for determining TSI in virally suppressed persons and for estimating HIV incidence in populations with high levels of viral suppression.

It will be important to test the performance of HIV-phyloTSI in different settings with different infection dynamics. If performance remains comparable to results obtained for the PopART Phylogenetics samples, the method could be used to measure HIV-1 incidence in a new population using a relatively small-scale phylogenetic cross-sectional survey. By sampling at least 1000 participants in an area with medium-level prevalence and incidence, it should soon be possible to estimate incidence using a mathematical model taking into account the fraction of HIV-positive participants, the fraction of viraemic participants and the TSI for viraemic participants. This approach for measuring incidence would require minimal data collection from participants and would be much less costly than measuring HIV incidence through repeated follow-up data collection in longitudinal cohorts.

In summary, HIV-phyloTSI is a powerful tool for estimating TSI, obtaining population-level recency estimates and estimating population-level incidence. Further work is required, but if performance is maintained in different settings, an improved HIV-phyloTSI tool has the potential to become a stepping stone in transforming HIV epidemiology in areas with generalised epidemics. This work would not have been possible without data contribution from multiple cohorts and consortia and highlights the importance of collaboration and data sharing in the area of HIV research.

## Data Availability

Model, example data, and code to obtain estimates from input data are freely available at https://github.com/BDI-pathogens/HIV-phyloTSI to facilitate application of HIV-phyloTSI to external datasets. Sequencing data and metadata are available by application to the PANGEA consortium, https://www.pangea-hiv.org.

## Abbreviations used

FRR: false recency rate
LRTT: longest root-to-tip distance
MAA: Multi-Assay Algorithms (for measuring TSI)
MAE: mean absolute error
MAF: minor allele frequency
MAF12c: MAF in the 1st and 2nd codons
MAF3c: MAF in the 3rd codon
MRCA: most recent common ancestor
NGS: next-generation sequencing
OLS: ordinary least squares (regression)
PrEP: Pre-Exposure Prophylaxis
TSI: time since infection (with HIV)
PreP: pre-exposure prophylaxis
UWP: University of Washington International Clinical Research Studies (ICRC) program datasets
RAK: Rakai Community Cohort Study dataset
BEE: BEEHIVE Consortium dataset
MRC: MRC-UVRI Uganda dataset

## Ethical considerations

All participants gave informed written consent, and ethics approvals were granted to the institutions that generated the data by in-country institutional review boards. In-country review board approvals were obtained from the following countries: Uganda for the Rakai Community Cohort study, the MRC-UVRI, the BEEHIVE collaboration and the UW cohorts, Zambia for the HPTN 071-02 PopART Phylogenetics study, Botswana, Kenya, South Africa and Tanzania for the UW cohorts and Belgium, Finland, France, Germany, the Netherlands, Sweden, Switzerland and the United Kingdom for the BEEHIVE cohorts. The design of the African studies was discussed and agreed with community advisory boards. The BEEHIVE study, which only accessed fully anonymised data, was approved by the ethics panel of the European Research Council. The University of Washington International Clinical Research Studies (ICRC) program was approved by IRB committees of the following institutions: University of Cape town, Research Ethics Committee; University of Witwaterstrand, HREC; Kenya Medical Research Institute; University of California San Francisco; Moi University; Indiana University; Kenyatta National Hospital; University of Washington; Kilimanjaro Christian Medical University College; London School of Hygiene and Tropical Medicine; Uganda National Council for Science and Technology (UNCST); Botswana Harvard Partnership (Republic of Botswana Ministry of Health, and Harvard School of Public Health). The Rakai Community Cohort study was approved by the Ugandan Virus Research Institute Scientific Research and Ethics Committee; the Ugandan National Council of Science and Technology; and the Western Institutional Review Board. The HPTN 071 (PopART) trial and the HPTN 071-02 (PopART) Phylogenetics Study was granted by ethics committees at the London School of Hygiene and Tropical Medicine, University of Zambia, and Stellenbosch University, South Africa.

## Funding statement

PANGEA is funded by the Bill & Melinda Gates Foundation (consecutive grants OPP1084362 and OPP1175094). BEEHIVE was funded by a European Research Council Advanced Grant (PBDR-339251). HPTN 071 is sponsored by the National Institute of Allergy and Infectious Diseases (NIAID) under Cooperative Agreements UM1-AI068619, UM1-AI068617, and UM1-AI068613, with funding from the US President’s Emergency Plan for AIDS Relief (PEPFAR). Additional funding is provided by the International Initiative for Impact Evaluation (3ie) with support from the Bill & Melinda Gates Foundation, as well as by the Division of Intramural Research, NIAID, the National Institute on Drug Abuse (NIDA), and the National Institute of Mental Health (NIMH), all part of NIH. HPTN 071-02 Phylogenetics is sponsored by NIAID, NIMH, and the Bill & Melinda Gates Foundation. The content is solely the responsibility of the authors and does not necessarily represent the official views of the NIAID, NIMH, NIDA, PEPFAR, 3ie, or the Bill & Melinda Gates Foundation. The computational aspects of this research were supported by the Wellcome Trust Core Award Grant Number 203141/Z/16/Z and the NIHR Oxford BRC. The funders had no role in study design, data collection and analysis, decision to publish, or preparation of the manuscript. The views expressed are those of the author(s) and not necessarily those of the NHS, the NIHR or the Department of Health.

## Author contributions

Conceptualization: TG, CF

Data curation: TG, LT

Formal Analysis: TG

Funding acquisition: CF, RH, HA, SF, MC

Investigation: DS, PK, MKG, RG, JK, JRL, JTH, NRM, AM, JB, CLC, ML, HA, BK, TdO, PK, DBo, GMC, DP, AG, FB, MB, DBe, SHO, JA, NB, JF, BGB, HFG, PKi, RDK, LM, KP, AVS, AVDV, BB, PKe, MC, PR

Methodology: TG, CF

Project administration: LAD, CW, CF, RH, HA, SF, ML

Resources: DS, PK, MKG, RG, JK, JRL, JTH, NRM, AM, JB, CLC, ML, HA, AG, FB, MB, DBe, SHO, JA, NB, JF, BGB, HFG, PKi, RDK, LM, KP, AVS, AVDV, BB, PKe, MC, PR

Software: TG Supervision: CF

Visualization: TG, MH, LAD, CF

Writing – original draft: LAD, TG, OL

Writing – review & editing: LAD, TG, OL, CF, MH, OL, OR, SE, RH, MC, CW

## Methods

### Sample collection and sequencing

Samples were collected from venous blood of HIV-1 viraemic individuals, and 0.5 ml of plasma was used for sequencing. All samples were sequenced using veSEQ-HIV (Bonsall et al. 2020) except samples from MRC Uganda/UVRI which were generated by pooling four overlapping PCR amplicons as described in Gall 2012 (Gall et al. 2012).

### Bioinformatic processing

Sequence reads were filtered to remove human and bacterial sequences using Kraken (Wood and Salzberg 2014), and assembled into contigs using SPAdes v3.10.1 (setting --meta) (Nurk et al. 2013). The resulting contigs were aligned to a curated alignment of 165 representative HIV genomes from the LANL HIV database (Kuiken, Korber, and Shafer 2003) to identify HIV contigs and generate a consensus sequence, followed by mapping the HIV reads onto the consensus. HIV genomes were assembled with shiver v1.3 (Wymant, Blanquart, et al. 2018), which generated a custom consensus sequence for each HIV sample. Shiver was run with Picard deduplication enabled (“Picard”), to eliminate duplicate reads that arise during the sequencing process. The final BAM files and base frequencies were coordinate-translated by shiver to bring them into alignment with the standard HXB2 HIV reference genome, RefSeq accession NC_001802. The reference alignment and primer sequences are available from the shiver GitHub repository (*Shiver: Sequences from HIV Easily Reconstructed*).

### Estimation of within-host diversity (MAF)

Within-host diversity was estimated as the cumulative minor allele frequency (MAF) at each genomic position:

MAF = (1 – proportion of majority base) / depth at each position, where depth was the number of unique (deduplicated) reads observed at that position. MAF values at first and second codon position were termed ‘MAF12c’, and MAF at third codon position was termed ‘MAF3c’.

### Estimation of within-host divergence (LRTT)

To estimate the extent of within-host divergence, each individual’s HIV sub-population was examined in a series of overlapping windows positioned every 10 bp along the entire length of the HIV genome, excluding the terminal repeat regions. For each 250 bp window, a maximum likelihood phylogeny was estimated using IQ-TREE (Nguyen et al. 2015) with the GTR+F+R6 model (generalised time reversible model with FreeRate with six categories of rate variation). Trees were processed in *phyloscanner* (Wymant, Hall, et al. 2018) with the settings -sks -ow -rda -swt 0.5 -amt -sat 0.33 -rcm -blr -pbk 15 -rtt 0.005 -rwt 3 -m 1E-5, and all statistics relating to the depth of the tree were extracted from the output of that package. Statistics related to tree depth and branch lengths were assessed for strength of correlation with the known time since infection (TSI) of the training data. These included LRTT, reported by phyloscanner as the field ‘normalised.largest.rtt’ within its patStats.csv output file; overall root-to-tip distance (‘normalised.overall.rtt’) and pairwise distance (‘subgraph.mean.patristic.distance’). The LRTT variable corresponds to the maximum evolutionary distance from the most recent common ancestor of the virus within the host, as observable in that window, normalised by genetic divergence at this genomic locus in the global HIV phylogeny. LRTT values at each window centre were collected for each sample. In addition, two other statistics that relate to the phylogenetic estimates were collected: ‘solo.dual.count’, representing the probability that the sample has come from a dual infection, and ‘tips’, representing the number of tips in the given window.

### Generation of aggregate statistics (feature engineering)

MAF3c, MAF12c and LRTT were calculated for the centre of each 250bp genomic window, averaged for each major gene (*gag, pol, gp120* and *gp41*) and across the whole genome. The mean number of tips was likewise calculated per gene and for the whole genome. A single genome-wide measure of number of windows supporting dual infection was generated by taking the mean of phyloscanner variable ‘solo.dual.count’ over all genomic windows, for each sample.

### Imputation of missing data

Where data were missing, for example because there were insufficient reads to determine the tree for that window, we tested two imputation strategies: zero-filling (on the assumption that lack of variation resulted in absence of an LRTT value), versus a more complex strategy of using the K-nearest neighbours (KNN) as implemented in TensorFlow, with the python package fancyimpute (Rubinsteyn). In order to enable robust calculation of Euclidean distances by fancyimpute, the features were first standardised to a mean of zero and standard deviation of 1. To guard against over-imputing, which can cause model over-fitting, we excluded windows where over 40% of samples had missing data, and excluded samples over 40% of windows had missing data. In addition, windows that included the position of the amplicon HIV-1 primers used for sequencing in the MRC cohort were excluded, as variation at these positions would be expected to be uninformative in the amplicon data. This resulted in a dataset containing 527 samples, with data at 820 genomic windows. We found that both imputation strategies had performed similarly on these samples, and chose KNN as the more robust method for the final model.

### Regression method selection

To select a regression method for predicting the midpoint time since infection (TSI), three different strategies were tested: ordinary least squares regression, gradient boosted regression, and random forest regression, as implemented in the scikit-learn package (Scikit-Learn). The midpoint is commonly used for estimating incidence, an alternative measure would have been a random-point estimate (Vandormael et al. 2018). Models were fitted using combinations of the MAF3c, MAF12c and LRTT aggregated predictors (Table S1). For each combination of predictors, the dataset was split into 75% training: 25% test data, and performance of each regression method was assessed as the r^2^ on the same fold of test data, for each combination of features. This procedure was repeated 10 times for cross-validation. Maximum depth of decision trees was constrained to 7 to prevent overfitting, and the random forest contained 1000 decision trees.

### Identification of most informative windows

The LRTT, MAF3c and MAF12c values within all genomic windows were individually assessed in univariate OLS regression models for prediction of TSI. The LRTT and MAF3c values at the most informative windows, defined as r^2^ of 0.3 or above, were used as one of the feature combinations for model selection (‘LRTT_MAF3c_topwin’).

### Feature selection

A table of possible feature combinations was generated for testing the predictive power of genome-wide values of MAF12c, MAF3c and LRTT, with or without the additional phylogenetic statistics of dual count and tip count. Feature combinations that used all or a subset of individual genomic windows were labelled as “FULL”, while aggregated features (averaged across each major HIV gene一*gag, pol, gp120* and *gp41*一and for the whole genome) were labelled “MEANS”. A binary indicator variable for the sequencing method (amplicons or veSeq) was always included. For each feature combination, predictive power was assessed with *k*-fold cross-validation. Random forest regression models, set up as described in ‘Regression method selection’, were trained on 75% of the data, leaving 25% as the test dataset; this procedure was repeated 20 times for cross-validation, each time assessing performance on the test dataset using the model r^2^ on test data, precision (width of 95% CI of the distribution of estimates returned from the 1000 decision trees within the forest), and mean absolute error (absolute difference between the expected and estimated TSI). Additionally, the expected accuracy and false recency rates were calculated, respectively, as the proportion of decision trees in the model that correctly returned estimates that fell below/above 1 year, and estimates below 1 year when the known midpoint TSI was above 1 year. Feature sets were preferred if they had a higher r^2^, accuracy and precision; where values were similar, the feature set with the lower expected false recency rate was chosen. The final feature set (designated ‘MEANS_feats_LRTT’) comprised: mean LRTT in each of *gag, pol* and *gp120*, MAF3c in *gag* and *gp41*, MAF12c in *gp41*, and the mean number of tips in each of *gag, gp41*, and *gp120*.

### TSI estimates for all samples

A leave-one-out strategy was used to obtain estimates of TSI for all samples, iterating over the full sample set, each time dropping the sample of interest, building a new random forest regression model on the remaining samples using the MEANS_feats_LRTT feature set, and obtaining a point estimate together with the full distribution of estimates from each of the 1000 decision trees in the forest. A separate random forest regression model was trained on the mean absolute errors from the base model, to generate 95% prediction intervals around the point estimates.

### Performance assessment by subtype

To check for evidence of subtype-dependent bias in model performance, we compared the mean absolute error (MAE) for all samples aggregated by subtype, after adjusting for TSI to account for the increase in MAE with increasing TSI. We fitted a linear model of MAE by TSI (MAE ∼ TSI) and compared the residuals by subtype, using Tukey’s range test for multiple comparison of means as implemented in the Python statsmodels library (statsmodels.stats.multicomp.pairwise_tukeyhsd), with alpha set to 0.05.

### Model performance in a simulated population

We simulated a population sample of 1000 HIV-infected individuals under the assumptions that the mean time to viral suppression in this population was 3 years. The duration of infection of individuals in this population with TSI *w* was modelled as 1 - e^-0.3w^. The predicted fraction of infections with estimated TSI up to *w* in this population, and the proportion of these that were incorrectly classified, were calculated using the false and true recency rates for the model at values of *w* between 3 and 36 months.

### Application to an independent dataset: PopART population cohort

The HPTN 071-02 Phylogenetics ancillary study to the HPTN 071 (PopART) trial collected samples from HIV-positive study participants in nine communities in Zambia between 2014 and 2019 (“HPTN 071-02 Study Protocol” 2017). Unused samples from vials collected to assess the main trial outcome were sequenced using veSeq-HIV. Sequences were assembled using *shiver*, and MAF values generated in the same way as for the training data. Sequences were batched in randomly allocated groups of 100 to build trees and obtain LRTT values, which were then used as inputs for the HIV-phyloTSI predictive model. Model outputs were TSI and estimated prediction interval. For the subset of 204 samples for which a last negative test date was available, midpoint predictions from HIV-phyloTSI were compared with midpoints of the known seroconversion interval, as was done for the training data.

## Supplement

**Figure S1:**
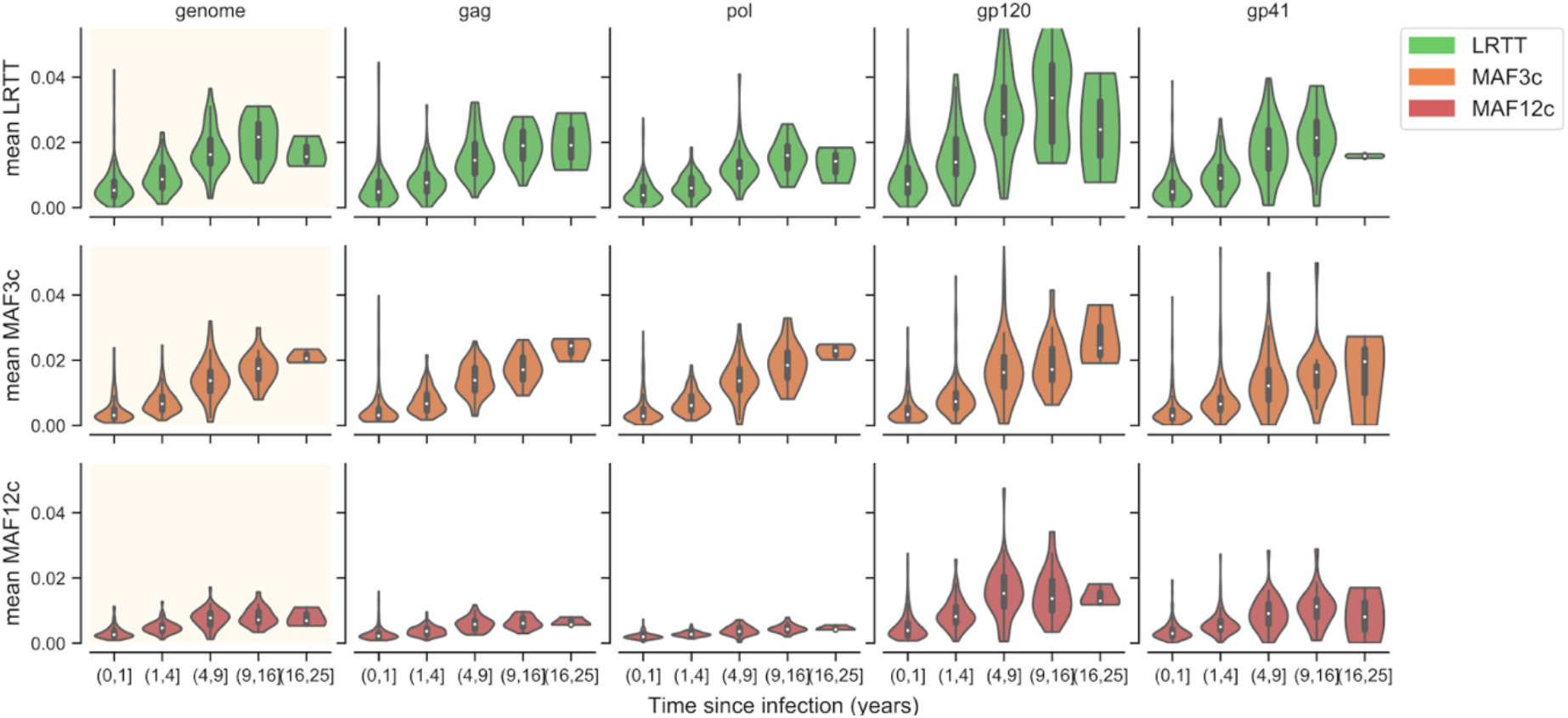
Genetic divergence (LRTT) and genetic diversity (MAF3c, MAF2c) for individual windows plateau at longer duration of infection. Vertical bars indicate 95% bootstrap confidence interval for the mean of each predictor, aggregated across the entire genome (first column, shaded) or within each of *gag, pol, gp120* and *gp41* HIV genes, for all samples.

**Figure S2.**
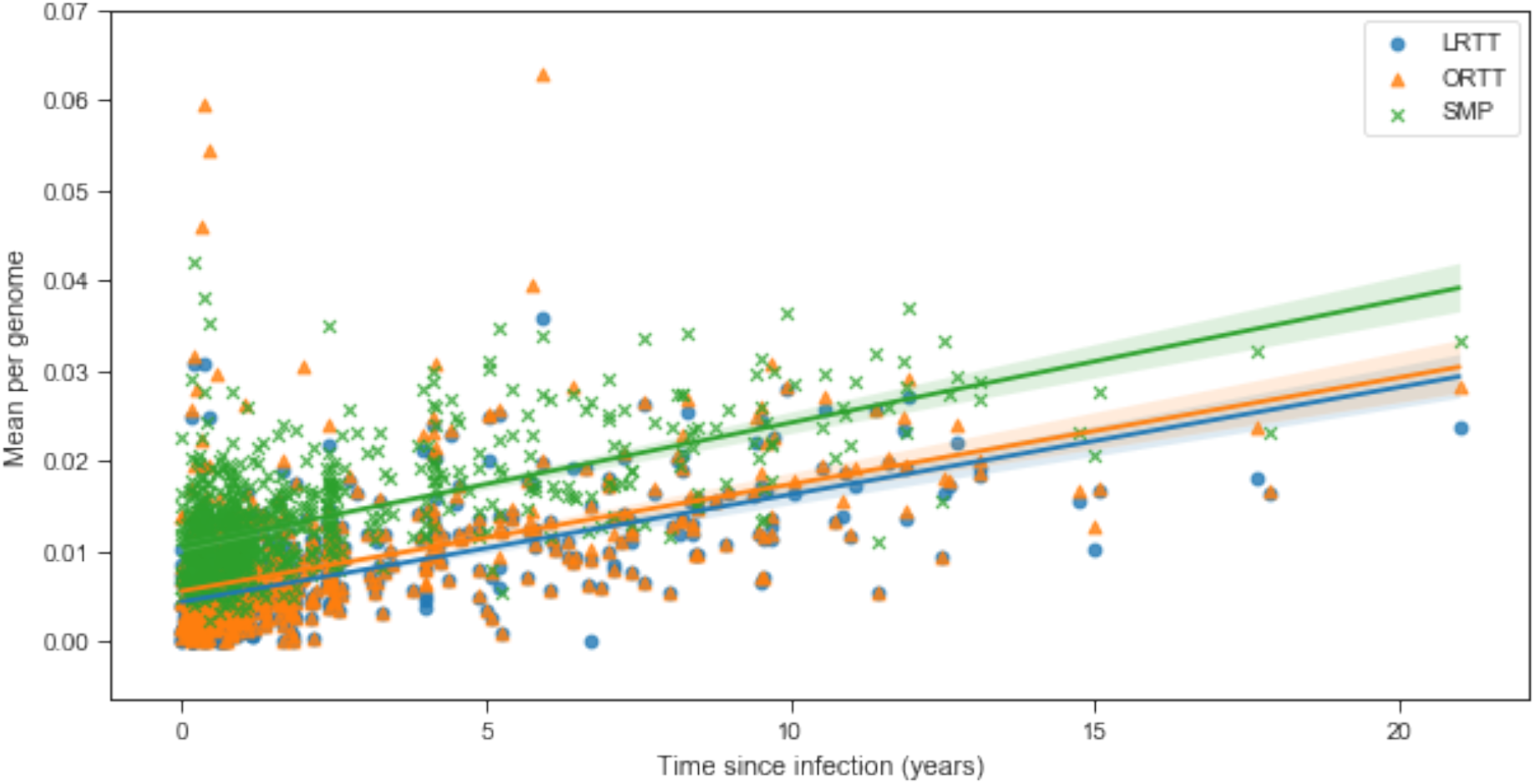
Phylogenetic metrics reported by *phyloscanner* are correlated with TSI. LRTT, largest subgraph root-to-tip distance; ORTT, overall root-to-tip distance; SMP, subgraph mean patristic distance. All three metrics are corrected with TSI, and are strongly correlated with one another, with some noisier estimates particularly for ORTT where a sample had multiple subgraphs due to e.g., contaminating reads in the window. For this analysis LLRT was selected, as this metric is most robust to presence of multiple subgraphs (i.e., individuals infected with multiple viral strains).

**Figure S3:**
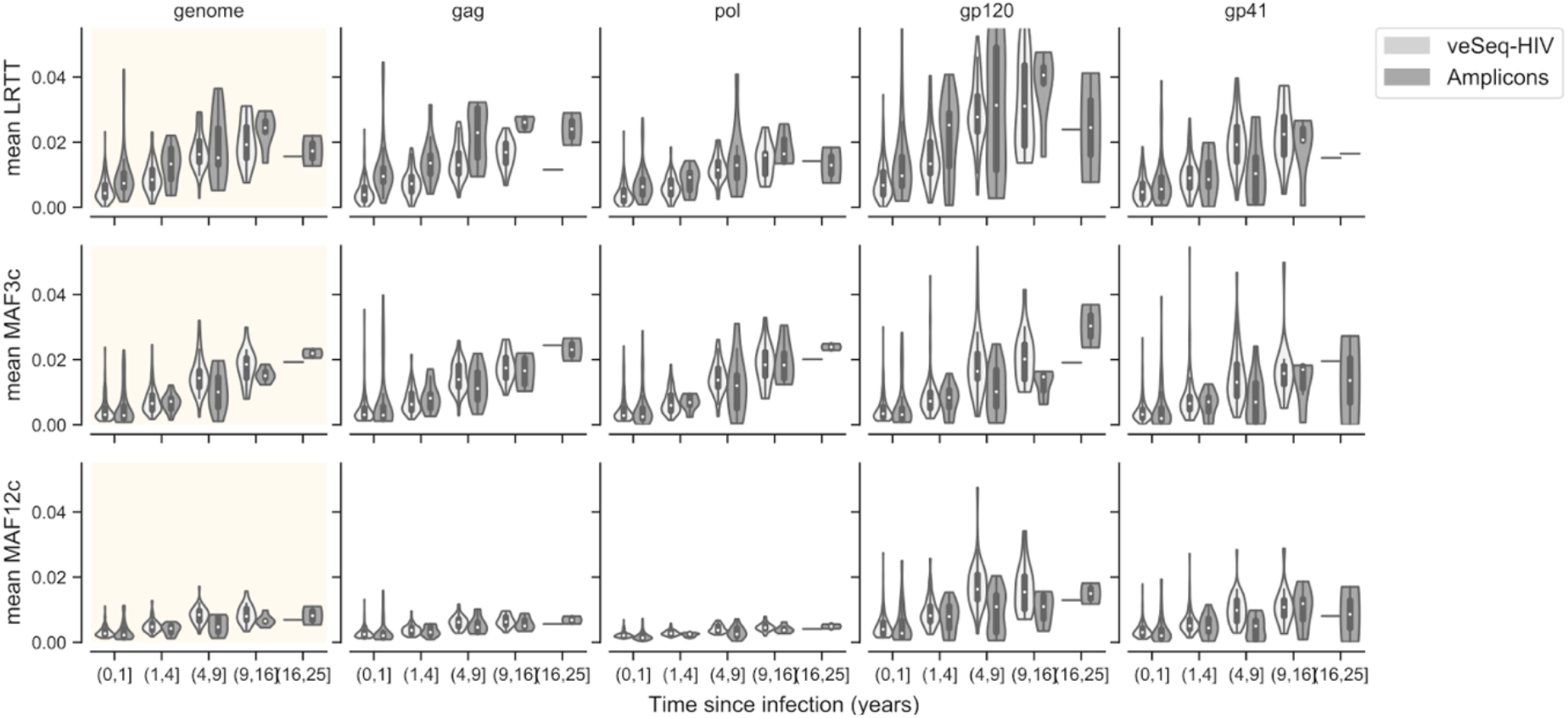
Mean divergence (LRTT) and diversity (MAF3c, MAF12c) measures increase with duration of infection in both veSeq-HIV and amplicon sequence data. Vertical bars indicate 95% bootstrap confidence interval for the mean of each predictor, aggregated across the entire genome (first column, shaded axes) or within each of gag, pol, gp120 and gp41 HIV genes, separately for veSeq-HIV sequences (light violins) and amplicon sequences (dark violins).

**Figure S4.**
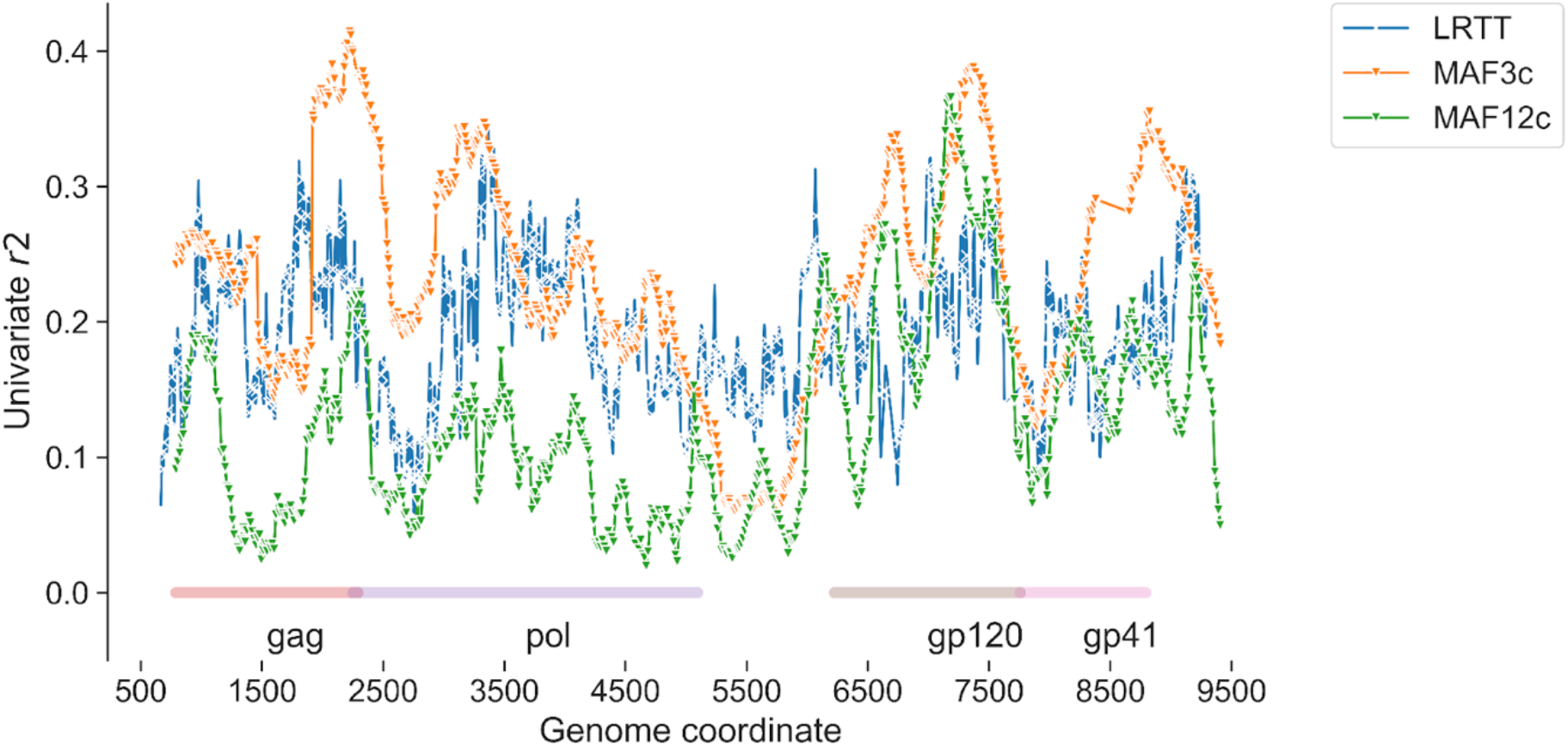
Correlations with TSI for predictors across the HIV-1 genome. Univariate linear regression models (OLS) were fitted independently for LRTT, MAF3c and MAF12c in overlapping 250b genomic windows, using the square root of the estimated duration of infection as the target variable. Missing data were zero-filled. Shown is the r2 within each window, with the window centre plotted on the x-axis.

**Table S1:** Cross-validated scores (mean r2 values in 10 folds) for different sets of LRTT, MAF3c and/or MAF12c feature combinations, computed using ordinary least squares (OLS), gradient boosted and random forest regression.

| Feature set | Predictors | Number of predictors | Regression Method |  |  |
| --- | --- | --- | --- | --- | --- |
|  |  |  | Linear OLS | Gradient boosted | Random forest |
| **genome_LRTT_MEAN** | is_mrc, genome_lrtt | 2 | 0.409 | 0.446 | 0.455 |
| **genome_MAF12c_MEAN** | is_mrc, genome_maf12c | 2 | 0.304 | 0.185 | 0.218 |
| **genome_MAF3c_MEAN** | is_mrc, genome_maf3c | 2 | 0.219 | 0.248 | 0.322 |
| **gag_LRTT_MEAN** | is_mrc, gag_lrtt | 2 | 0.377 | 0.412 | 0.412 |
| **gag_MAF3c_MEAN** | is_mrc, gag_maf3c | 2 | -0.038 | 0.388 | 0.452 |
| **gp120_LRTT_MEAN** | is_mrc, gp120_lrtt | 2 | 0.335 | 0.312 | 0.323 |
| **gp120_MAF3c_MEAN** | is_mrc, gp120_maf3c | 2 | 0.252 | 0.341 | 0.349 |
| **gp41_LRTT_MEAN** | is_mrc, gp41_lrtt | 2 | 0.294 | 0.221 | 0.229 |
| **gp41_MAF3c_MEAN** | is_mrc, gp41_maf3c | 2 | 0.107 | 0.211 | 0.226 |
| **pol_LRTT_MEAN** | is_mrc, pol_lrtt | 2 | 0.334 | 0.290 | 0.302 |
| **pol_MAF3c_MEAN** | is_mrc, pol_maf3c | 2 | 0.051 | 0.291 | 0.310 |
| **genome_MAF_MEANS** | is_mrc, genome_maf3c, genome_maf12c | 3 | 0.206 | 0.248 | 0.329 |
| **LRTT_MEANS** | is_mrc, gag_lrtt, pol_lrtt, gp120_lrtt, gp41_lrtt, genome_lrtt | 6 | 0.406 | 0.517 | 0.558 |
| **MAF12c_MEANS** | is_mrc, gag_maf12c, pol_maf12c, gp120_maf12c, gp41_maf12c, genome_maf12c | 6 | 0.267 | 0.375 | 0.434 |
| **MAF3c_MEANS** | is_mrc, gag_maf3c, pol_maf3c, gp120_maf3c, gp41_maf3c, genome_maf3c | 6 | 0.060 | 0.449 | 0.463 |
| **genome_LRTT_MAF_MEANS** | is_mrc, genome_tips, genome_dual, genome_lrtt, genome_maf3c,<br>genome_maf12c | 6 | 0.522 | 0.500 | 0.532 |
| **genome_MEANS** | is_mrc, genome_tips,<br>genome_dual,<br>genome_lrtt,<br>genome_maf3c,<br>genome_maf12c | 6 | 0.522 | 0.500 | 0.532 |
| **MEANS_feats** | is_mrc, genome_lrtt,<br>genome_maf3c, gag_lrtt,<br>gag_maf3c, pol_lrtt,<br>gp41_lrtt, pol_maf12c | 8 | 0.383 | 0.594 | 0.598 |
| **MEANS_feats_LRTT** | is_mrc, gag_lrtt, pol_lrtt,<br>gp120_lrtt, gag_maf3c,<br>gp41_maf3c,<br>gp41_maf12c, gag_tips,<br>gp41_tips, gp120_tips | 10 | 0.489 | 0.640 | 0.650 |
| **MEANS_notips_nodual** | is_mrc, gag_lrtt,<br>gag_maf3c, gag_maf12c,<br>pol_lrtt, pol_maf3c,<br>pol_maf12c, gp120_lrtt,<br>gp120_maf3c,<br>gp120_maf12c,<br>gp41_lrtt, gp41_maf3c,<br>gp41_maf12c,<br>genome_lrtt,<br>genome_maf3c,<br>genome_maf12c | 16 | 0.320 | 0.614 | 0.630 |
| **GPE_MEANS** | is_mrc, genome_tips,<br>genome_dual, gag_lrtt,<br>gag_tips, gag_maf3c,<br>gag_maf12c, pol_lrtt,<br>pol_tips, pol_maf3c,<br>pol_maf12c, gp120_lrtt,<br>gp120_tips,<br>gp120_maf3c,<br>gp120_maf12c,<br>gp41_lrtt, gp41_tips,<br>gp41_maf3c,<br>gp41_maf12c | 19 | 0.452 | 0.636 | 0.638 |

**Figure S5.**
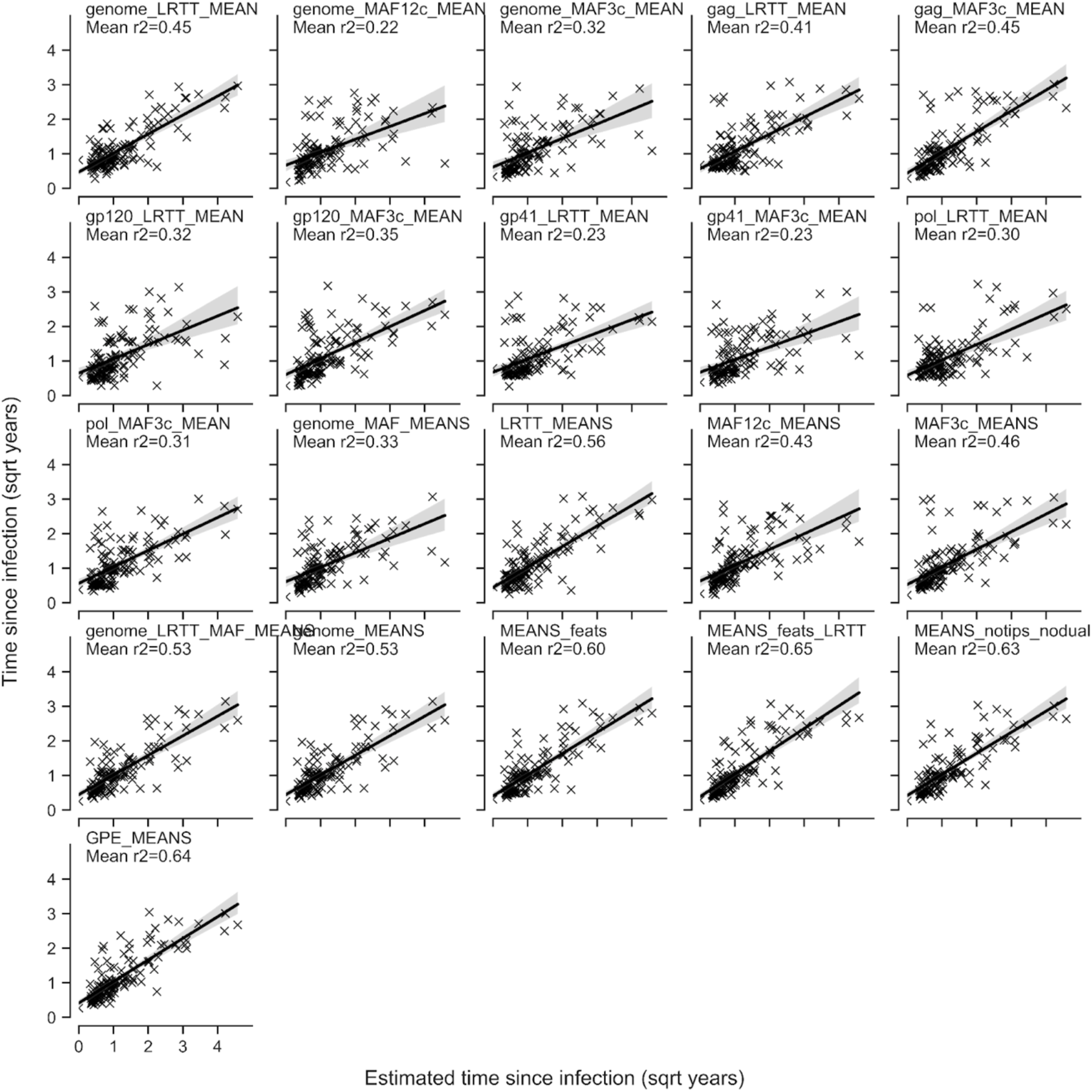
Scatterplots of known versus estimated square root-transformed time since infection for aggregate regression models in Table S2, derived using random forest regression. Models are named in correspondence with Table S2. Shown are scatterplots for the same set of training:test data (fold) for all models.

**Figure S6.**
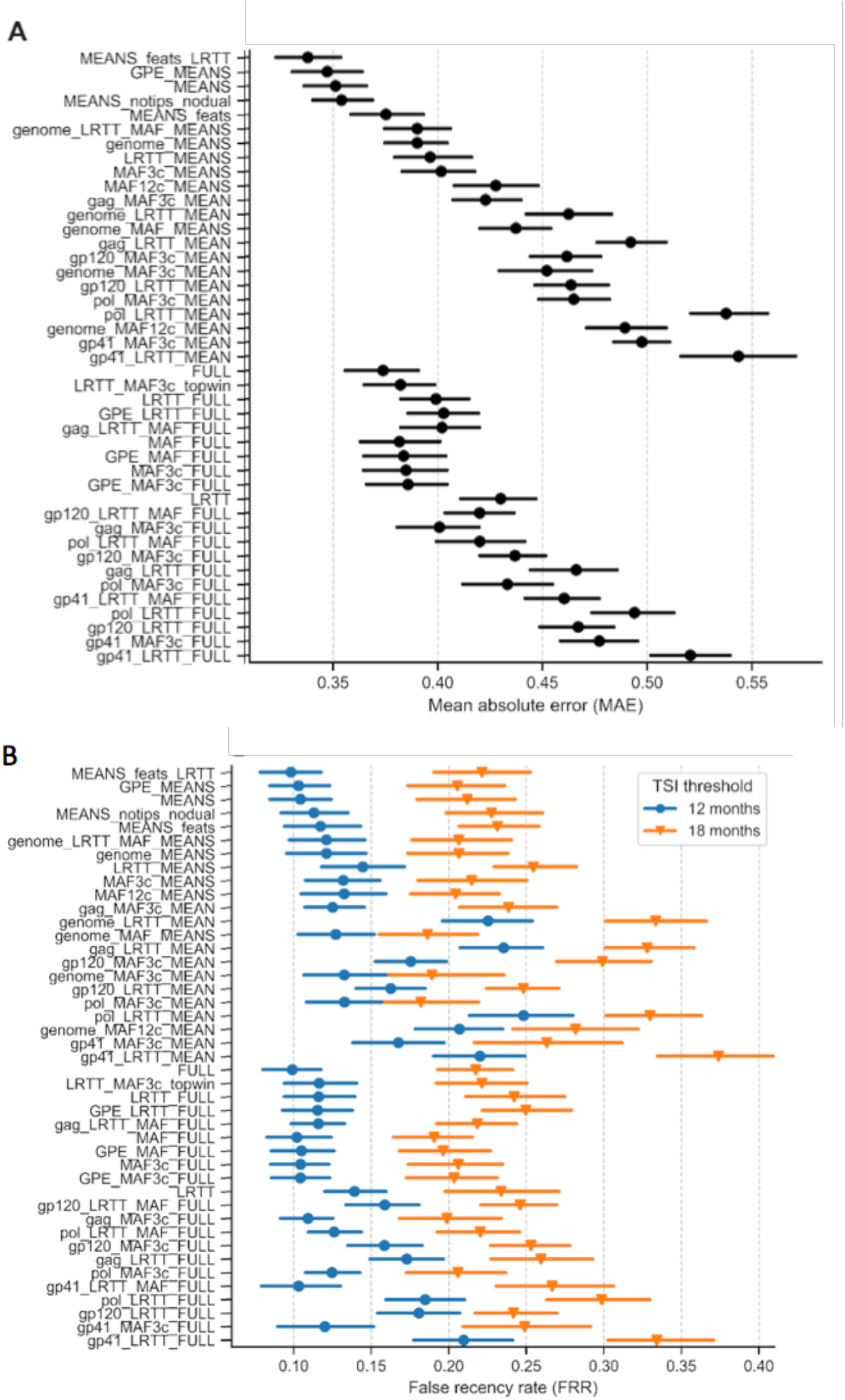
Mean absolute error (MAE) and estimated false recency rate (FRR). In all panels, circles show mean and lines show 95% bootstrap CI over 20-fold cross-validation. A. MAE, calculated as absolute difference between known and estimated TSI in square-root space. B. D. FRR, computed as the fraction of samples with known TSI over 12 months (circles, blue) or 18 months (triangles, orange) for which TSI was incorrectly estimated as being below 12 or 18 months, respectively.

**Figure S7.**
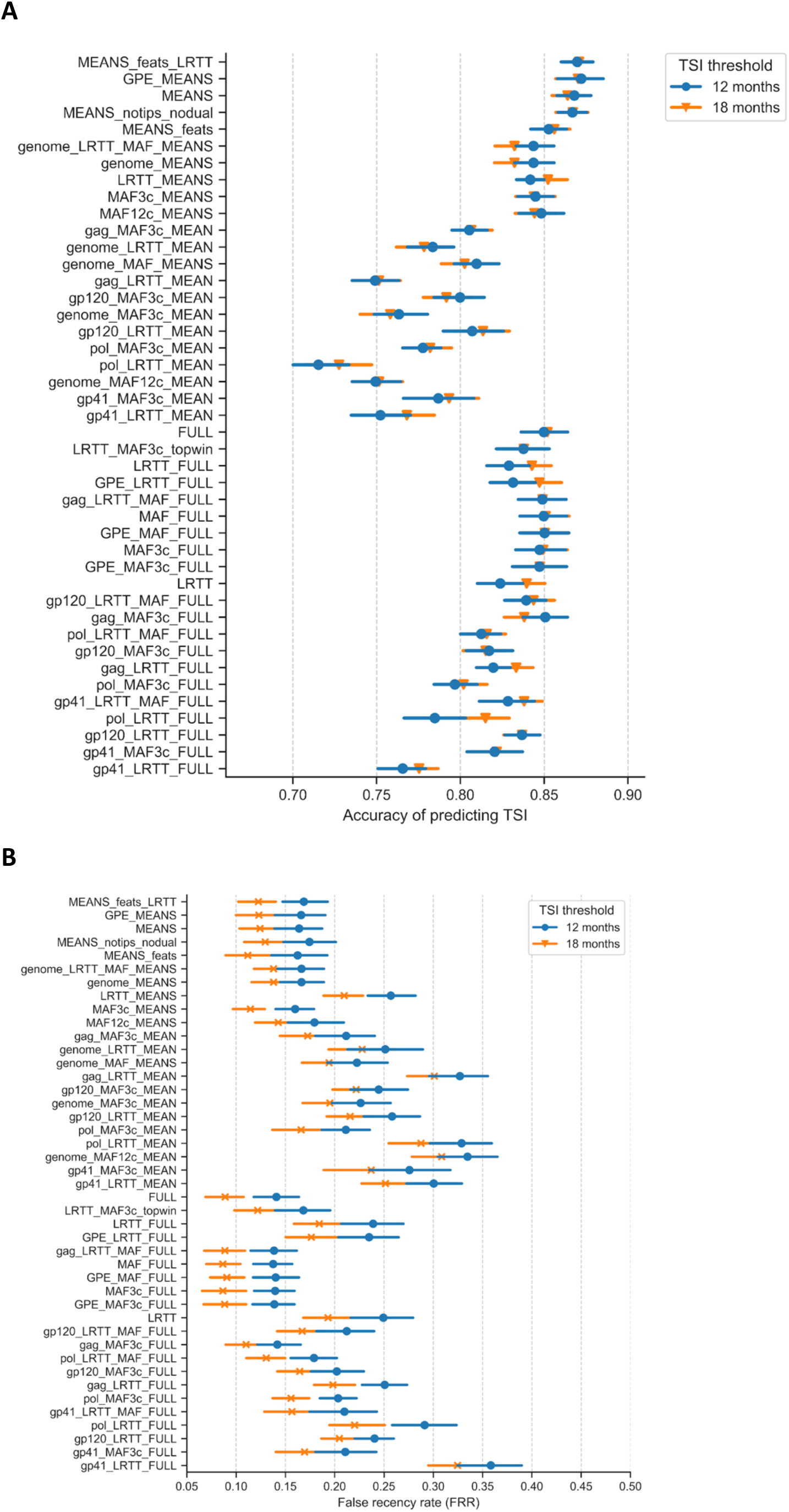
Accuracy and False Recency Rate (FRR) of classification models. Points show mean score; vertical lines indicate 95% CI over 20-fold cross-validation. A. Accuracy was calculated as the proportion of samples correctly predicted at a threshold of 12 months (blue circles) or 18 months (orange triangles), out of the total number of samples. B. False recency rate was calculated as the proportion of non-recent samples (with TSI above threshold) incorrectly classified as being recent with TSI < threshold at either 12 months (blue circles) or 18 months (orange crosses).

**Figure S8.**
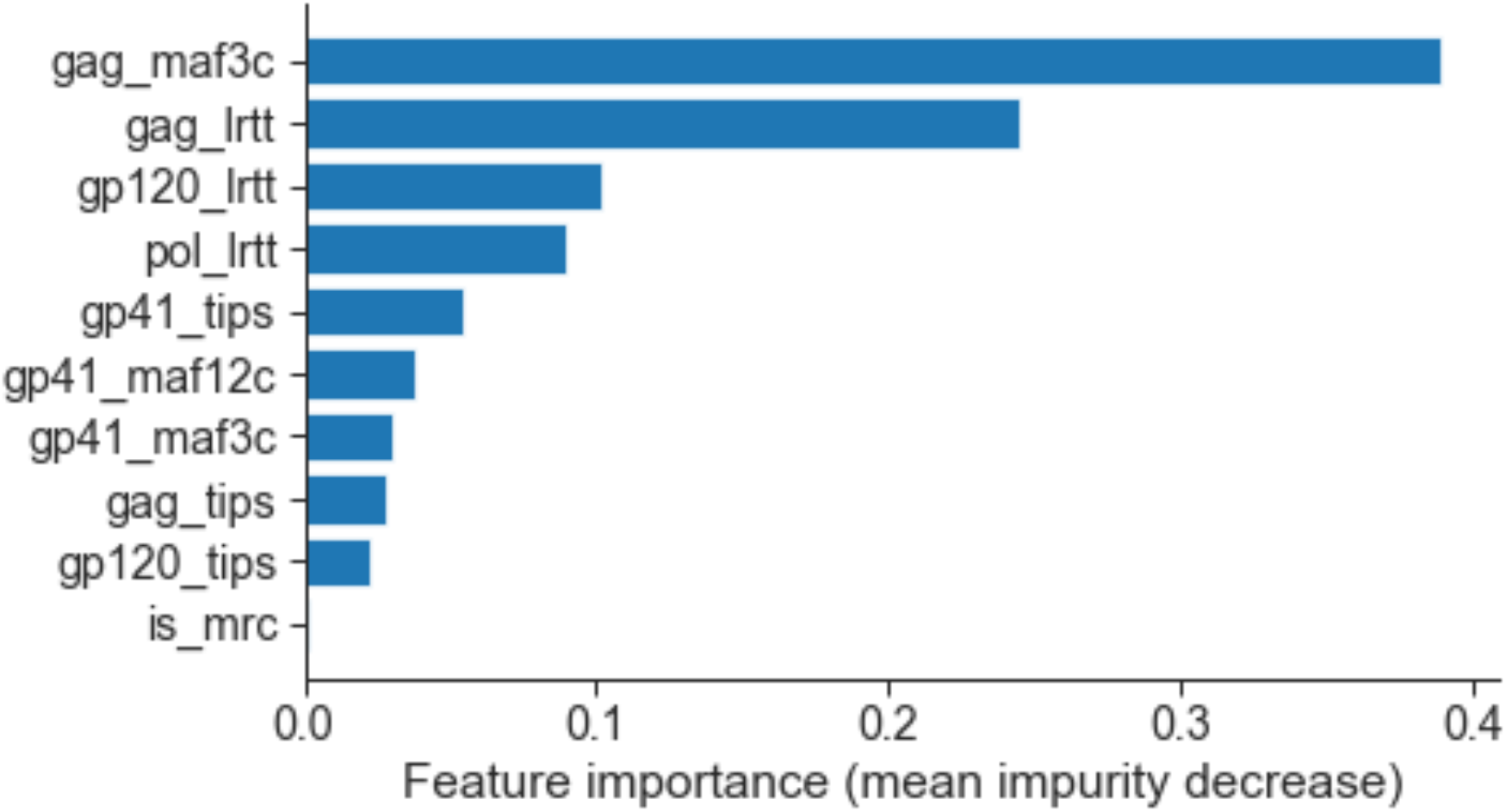
Feature importances (mean decrease in impurity among 1000 decision trees) for feature set MEANS_feats_LRTT. Contribution of each feature (proportional to variance explained) for the ten features in the best-performing regression models. Feature importances were extracted from the random forest model feature_importances_ attribute, computed within scikit-learn as the mean of accumulation of the impurity decrease within each tree.

**Figure S9.**
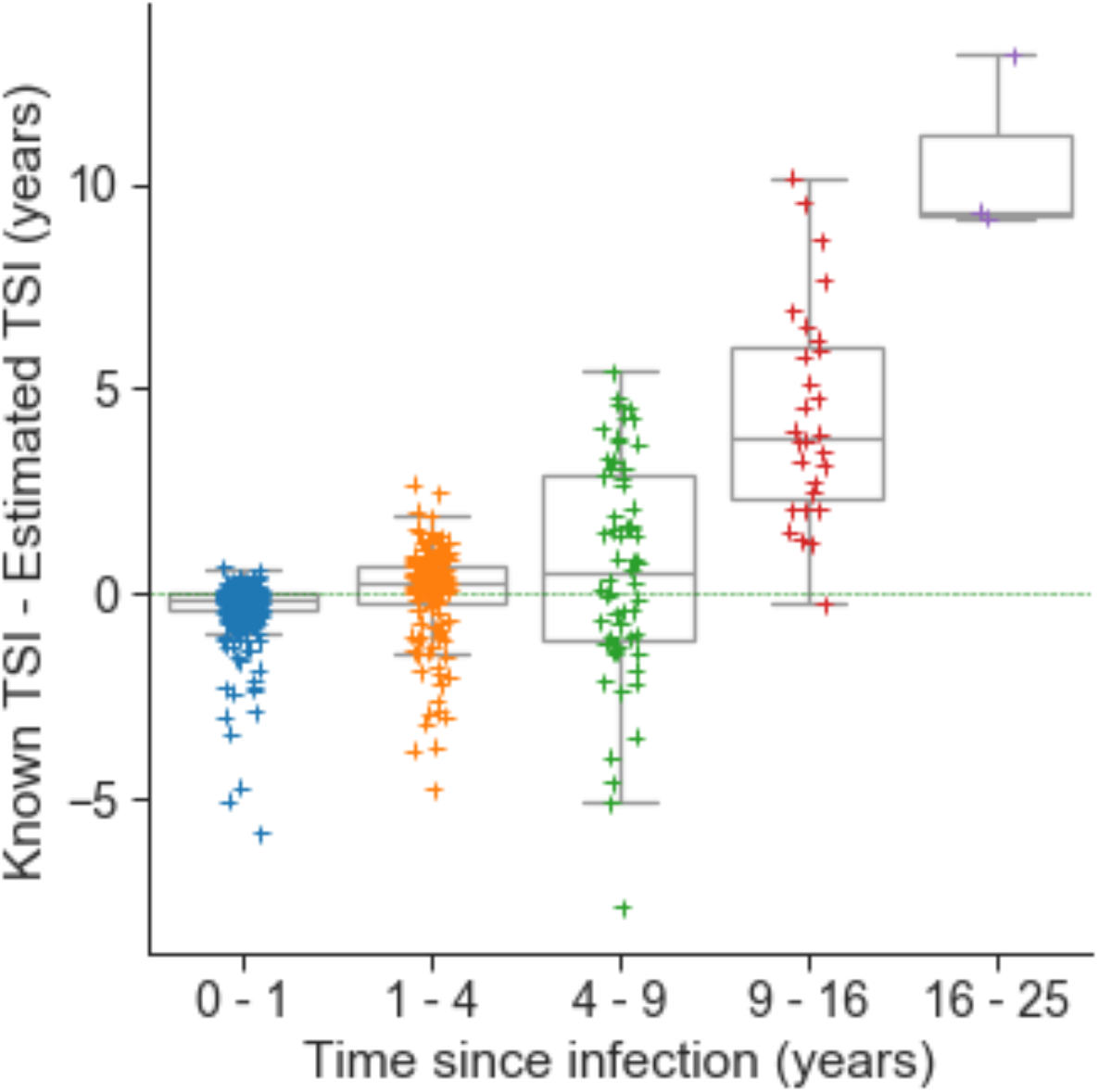
Model bias is low for infections <9 years. Boxplots show the median and quartiles for each TSI category; whiskers extend to 1.5 IQR. The difference between known and estimated TSI increases for non-recent infections. TSI for recent infections (under 1 year) tends to be slightly overestimated, while TSI for long-term infections is increasingly underestimated.

**Table S2.** Accuracy, false recency rate and true recency rate for simulated population data, with recency defined variously as infections occurring in the preceding 3, 6, 12, 18 and 24 months. One thousand individuals were drawn from a population with an average interval of three years from infection to treatment.

| Months | Accuracy | False recency rate | True recency rate |
| --- | --- | --- | --- |
| 3 | 93.0 | 2.2 | 49.7 |
| 6 | 86.6 | 4.7 | 65.9 |
| 12 | 80.1 | 11.6 | 74.0 |
| 18 | 86.1 | 10.3 | 84.6 |
| 24 | 89.6 | 5.8 | 88.4 |

**Table S3.** Tukey range test result for pairwise subtype comparisons of model bias. Mean bias of model predictions for all samples, adjusted for time since infection, compared using the Tukey range test for pairwise comparison of means, as implemented in the statsmodels python library. None of the subtypes differed at p<0.05 (reject=False).

| Group 1 | Group 2 | Mean between-group difference | Lower | Upper | Reject<br>($p < 0.05$) |
| --- | --- | --- | --- | --- | --- |
| A1 | B | -0.0079 | -0.1575 | 0.1417 | FALSE |
| A1 | C | 0.0188 | -0.104 | 0.1416 | FALSE |
| A1 | D | -0.0015 | -0.1369 | 0.1339 | FALSE |
| A1 | Other | -0.0251 | -0.17 | 0.1198 | FALSE |
| B | C | 0.0267 | -0.1095 | 0.1629 | FALSE |
| B | D | 0.0064 | -0.1413 | 0.154 | FALSE |
| B | Other | -0.0171 | -0.1736 | 0.1393 | FALSE |
| C | D | -0.0203 | -0.1407 | 0.1001 | FALSE |
| C | Other | -0.0438 | -0.1749 | 0.0872 | FALSE |
| D | Other | -0.0235 | -0.1664 | 0.1194 | FALSE |

## Notes

### Competing Interest Statement

The authors have declared no competing interest.

## References

Abeler-Dörner, Lucie, Mary K. Grabowski, Andrew Rambaut, Deenan Pillay, Christophe Fraser, and PANGEA consortium. 2019. “PANGEA-HIV 2: Phylogenetics And Networks for Generalised Epidemics in Africa.” Current Opinion in HIV and AIDS 14 (3): 173–80.

Asiki, Gershim, Juliet Mpendo, Andrew Abaasa, Collins Agaba, Annet Nanvubya, Leslie Nielsen, Janet Seeley, Pontiano Kaleebu, Heiner Grosskurth, and Anatoli Kamali. 2011. “HIV and Syphilis Prevalence and Associated Risk Factors among Fishing Communities of Lake Victoria, Uganda.” Sexually Transmitted Infections 87 (6): 511–15.

Asiki, Gershim, Georgina Murphy, Jessica Nakiyingi-Miiro, Janet Seeley, Rebecca N. Nsubuga, Alex Karabarinde, Laban Waswa, et al. 2013. “The General Population Cohort in Rural South-Western Uganda: A Platform for Communicable and Non-Communicable Disease Studies.” International Journal of Epidemiology 42 (1): 129–41.

Baeten, Jared M., Deborah Donnell, Patrick Ndase, Nelly R. Mugo, James D. Campbell, Jonathan Wangisi, Jordan W. Tappero, et al. 2012. “Antiretroviral Prophylaxis for HIV Prevention in Heterosexual Men and Women.” The New England Journal of Medicine 367 (5): 399–410.

Blanquart, François, Chris Wymant, Marion Cornelissen, Astrid Gall, Margreet Bakker, Daniela Bezemer, Matthew Hall, et al. 2017. “Viral Genetic Variation Accounts for a Third of Variability in HIV-1 Set-Point Viral Load in Europe.” PLoS Biology 15 (6): e2001855.

Bonsall, David, Tanya Golubchik, Mariateresa de Cesare, Mohammed Limbada, Barry Kosloff, George MacIntyre-Cockett, Matthew Hall, et al. 2020. “A Comprehensive Genomics Solution for HIV Surveillance and Clinical Monitoring in Low-Income Settings.” Journal of Clinical Microbiology 58 (10). https://doi.org/10.1128/JCM.00382-20.

Brookmeyer, R. 1997. “Accounting for Follow-up Bias in Estimation of Human Immunodeficiency Virus Incidence Rates.” Journal of the Royal Statistical Society. Series A, 160 (1): 127–40.

Busch, Michael P., Christopher D. Pilcher, Timothy D. Mastro, John Kaldor, Gaby Vercauteren, William Rodriguez, Christine Rousseau, et al. 2010. “Beyond Detuning: 10 Years of Progress and New Challenges in the Development and Application of Assays for HIV Incidence Estimation.” AIDS 24 (18): 2763–71.

Carlisle, Louisa A., Teja Turk, Katharina Kusejko, Karin J. Metzner, Christine Leemann, Corinne D. Schenkel, Nadine Bachmann, et al. 2019. “Viral Diversity Based on Next-Generation Sequencing of HIV-1 Provides Precise Estimates of Infection Recency and Time Since Infection.” The Journal of Infectious Diseases 220 (2): 254–65.

Celum, C., A. Wald, J. R. Lingappa, A. S. Magaret, R. S. Wang, N. Mugo, A. Mujugira, et al. 2010. “Acyclovir and Transmission of HIV-1 from Persons Infected with HIV-1 and HSV-2.” The New England Journal of Medicine 362 (5): 427–39.

Chang, Larry W., Mary K. Grabowski, Robert Ssekubugu, Fred Nalugoda, Godfrey Kigozi, Betty Nantume, Justin Lessler, et al. 2016. “Heterogeneity of the HIV Epidemic in Agrarian, Trading, and Fishing Communities in Rakai, Uganda: An Observational Epidemiological Study.” The Lancet. HIV 3 (8): e388–96.

Cousins, Matthew M., Jacob Konikoff, Oliver Laeyendecker, Connie Celum, Susan P. Buchbinder, George R. Seage 3rd, Gregory D. Kirk, et al. 2014. “HIV Diversity as a Biomarker for HIV Incidence Estimation: Including a High-Resolution Melting Diversity Assay in a Multiassay Algorithm.” Journal of Clinical Microbiology 52 (1): 115–21.

Fidler, Sarah, Wolfgang Stöhr, Matt Pace, Lucy Dorrell, Andrew Lever, Sarah Pett, Sabine Kinloch-de Loes, et al. 2020. “Antiretroviral Therapy Alone versus Antiretroviral Therapy with a Kick and Kill Approach, on Measures of the HIV Reservoir in Participants with Recent HIV Infection (the RIVER Trial): A Phase 2, Randomised Trial.” The Lancet 395 (10227): 888–98.

Fogel, Jessica M., David Bonsall, Vanessa Cummings, Rory Bowden, Tanya Golubchik, Mariateresa de Cesare, Ethan A. Wilson, et al. 2020. “Performance of a High-Throughput next-Generation Sequencing Method for Analysis of HIV Drug Resistance and Viral Load.” The Journal of Antimicrobial Chemotherapy 75 (12): 3510–16.

Gall, Astrid, Bridget Ferns, Clare Morris, Simon Watson, Matthew Cotten, Mark Robinson, Neil Berry, Deenan Pillay, and Paul Kellam. 2012. “Universal Amplification, next-Generation Sequencing, and Assembly of HIV-1 Genomes.” Journal of Clinical Microbiology 50 (12): 3838–44.

Hall, Matthew, Tanya Golubchik, David Bonsall, Lucie Abeler-Dörner, Mohammed Limbada, Barry Kosloff, Ab Schaap, et al. 2021. “Demographic Characteristics of Sources of HIV-1 Transmission in Zambia.” bioRxiv. https://doi.org/10.1101/2021.10.04.21263560.

Hayes, Richard J., Deborah Donnell, Sian Floyd, Nomtha Mandla, Justin Bwalya, Kalpana Sabapathy, Blia Yang, et al. 2019. “Effect of Universal Testing and Treatment on HIV Incidence - HPTN 071 (PopART).” The New England Journal of Medicine 381 (3): 207–18.

HIV-phyloTSI: Estimate Time since Infection (TSI) from HIV Deep-Sequencing Data. https://github.com/BDI-pathogens/HIV-phyloTSI.

HPTN 071-02 Study Protocol 2017. https://www.hptn.org/sites/default/files/inline-files/HPTN%20071-2%2C%20Version%202.0%20%2807-14-2017%29.pdf.

Incidence Assay Critical Path Working Group. 2011. “More and Better Information to Tackle HIV Epidemics: Towards Improved HIV Incidence Assays.” PLoS Medicine 8 (6): e1001045.

Jain, Vivek, Wendy Hartogensis, Peter Bacchetti, Peter W. Hunt, Hiroyu Hatano, Elizabeth Sinclair, Lorrie Epling, et al. 2013. “Antiretroviral Therapy Initiated Within 6 Months of HIV Infection Is Associated With Lower T-Cell Activation and Smaller HIV Reservoir Size.” The Journal of Infectious Diseases. https://doi.org/10.1093/infdis/jit311.

Kasamba, Ivan, Stephen Nash, Maryam Shahmanesh, Kathy Baisley, Jim Todd, Onesmus Kamacooko, Yunia Mayanja, Janet Seeley, and Helen A. Weiss. 2019. “Missed Study Visits and Subsequent HIV Incidence Among Women in a Predominantly Sex Worker Cohort Attending a Dedicated Clinic Service in Kampala, Uganda.” JAIDS Journal of Acquired Immune Deficiency Syndromes. https://doi.org/10.1097/qai.0000000000002143.

Kouyos, Roger D., Viktor von Wyl, Sabine Yerly, Jürg Böni, Philip Rieder, Beda Joos, Patrick Taffé, et al. 2011. “Ambiguous Nucleotide Calls from Population-Based Sequencing of HIV-1 Are a Marker for Viral Diversity and the Age of Infection.” Clinical Infectious Diseases: An Official Publication of the Infectious Diseases Society of America 52 (4): 532–39.

Kuiken, Carla, Bette Korber, and Robert W. Shafer. 2003. “HIV Sequence Databases.” AIDS Reviews 5 (1): 52–61.

Lingappa, Jairam R., Slavé Petrovski, Erin Kahle, Jacques Fellay, Kevin Shianna, M. Juliana McElrath, Katherine K. Thomas, et al. 2011. “Genomewide Association Study for Determinants of HIV-1 Acquisition and Viral Set Point in HIV-1 Serodiscordant Couples with Quantified Virus Exposure.” PloS One 6 (12): e28632.

Longosz, Andrew F., Charles S. Morrison, Pai-Lien Chen, Eric Arts, Immaculate Nankya, Robert A. Salata, Veronica Franco, Thomas C. Quinn, Susan H. Eshleman, and Oliver Laeyendecker. 2014. “Immune Responses in Ugandan Women Infected with Subtypes A and D HIV Using the BED Capture Immunoassay and an Antibody Avidity Assay.” Journal of Acquired Immune Deficiency Syndromes 65 (4): 390–96.

Lundgren, Erik, Ethan Romero-Severson, Jan Albert, and Thomas Leitner. 2021. “Combining Biomarker and Virus Phylogenetic Models Improves Epidemiological Source Identification.” bioRxiv. https://doi.org/10.1101/2021.12.13.472340.

Mastro, Timothy D. 2013. “Determining HIV Incidence in Populations: Moving in the Right Direction.” The Journal of Infectious Diseases.

Moyo, Sikhulile, Alain Vandormael, Eduan Wilkinson, Susan Engelbrecht, Simani Gaseitsiwe, Kenanao P. Kotokwe, Rosemary Musonda, et al. 2016. “Analysis of Viral Diversity in Relation to the Recency of HIV-1C Infection in Botswana.” PloS One 11 (8): e0160649.

Moyo, Sikhulile, Eduan Wilkinson, Alain Vandormael, Rui Wang, Jia Weng, Kenanao P. Kotokwe, Simani Gaseitsiwe, et al. 2017. “Pairwise Diversity and tMRCA as Potential Markers for HIV Infection Recency.” Medicine 96 (6): e6041.

Nguyen, Lam-Tung, Heiko A. Schmidt, Arndt von Haeseler, and Bui Quang Minh. 2015. “IQ-TREE: A Fast and Effective Stochastic Algorithm for Estimating Maximum-Likelihood Phylogenies.” Molecular Biology and Evolution 32 (1): 268–74.

Nurk, Sergey, Anton Bankevich, Dmitry Antipov, Alexey A. Gurevich, Anton Korobeynikov, Alla Lapidus, Andrey D. Prjibelski, et al. 2013. “Assembling Single-Cell Genomes and Mini-Metagenomes from Chimeric MDA Products.” Journal of Computational Biology: A Journal of Computational Molecular Cell Biology 20 (10): 714–37.

Park, Sung Yong, Tanzy M. T. Love, Jeremy Nelson, Sally W. Thurston, Alan S. Perelson, and Ha Youn Lee. 2011. “Designing a Genome-Based HIV Incidence Assay with High Sensitivity and Specificity.” AIDS 25 (16): F13–19. Picard. https://broadinstitute.github.io/picard/.

Puller, Vadim, Richard Neher, and Jan Albert. 2017. “Estimating Time of HIV-1 Infection from next-Generation Sequence Diversity.” PLoS Computational Biology 13 (10): e1005775.

Ragonnet-Cronin, Manon, Stéphane Aris-Brosou, Isabelle Joanisse, Harriet Merks, Dominic Vallée, Kyna Caminiti, Michael Rekart, et al. 2012. “Genetic Diversity as a Marker for Timing Infection in HIV-Infected Patients: Evaluation of a 6-Month Window and Comparison with BED.” The Journal of Infectious Diseases 206 (5): 756–64.

Ragonnet-Cronin, Manon, Tanya Golubchik, Sikhulile Moyo, Christophe Fraser, Max Essex, Vlad Novitsky, Erik Volz, and with the PANGEA Consortium. 2021. “HIV Genetic Diversity Informs Stage of HIV-1 Infection among Patients Receiving Antiretroviral Therapy in Botswana.” The Journal of Infectious Diseases, June. https://doi.org/10.1093/infdis/jiab293.

Richman, Douglas D., David M. Margolis, Martin Delaney, Warner C. Greene, Daria Hazuda, and Roger J. Pomerantz. 2009. “The Challenge of Finding a Cure for HIV Infection.” Science 323 (5919): 1304–7.

Rubinsteyn, Alex. Fancyimpute: Multivariate Imputation and Matrix Completion Algorithms Implemented in Python. Github. Accessed November 23, 2021. https://github.com/iskandr/fancyimpute.

Ruone, Susan, Lynn Paxton, Tony McLaurin, Allan Taylor, Debra Hanson, Walid Heneine, John T. Brooks, and José Gerardo García-Lerma. 2016. “Brief Report: HIV-1 Evolution in Breakthrough Infections in a Human Trial of Oral Pre-Exposure Prophylaxis with Emtricitabine and Tenofovir Disoproxil Fumarate.” Journal of Acquired Immune Deficiency Syndromes 72 (2): 129–32.

Saito, Suzue, Yen T. Duong, Melissa Metz, Kiwon Lee, Hetal Patel, Katrina Sleeman, Julius Manjengwa, et al. 2017. “Returning HIV-1 Viral Load Results to Participant-Selected Health Facilities in National Population-Based HIV Impact Assessment (PHIA) Household Surveys in Three Sub-Saharan African Countries, 2015 to 2016.” Journal of the International AIDS Society 20 Suppl 7 (November). https://doi.org/10.1002/jia2.25004.

Scikit-Learn: Machine Learning in Python — Scikit-Learn 0.16.1 Documentation. http://scikit-learn.org/.

Shankarappa, R., J. B. Margolick, S. J. Gange, A. G. Rodrigo, D. Upchurch, H. Farzadegan, P. Gupta, et al. 1999. “Consistent Viral Evolutionary Changes Associated with the Progression of Human Immunodeficiency Virus Type 1 Infection.” Journal of Virology 73 (12): 10489–502.

Shiver: Sequences from HIV Easily Reconstructed. https://github.com/ChrisHIV/shiver.

Teixeira, Sylvia Lm, Cristina M. Jalil, Emilia M. Jalil, Sandro C. Nazer, Simone da Costa Cruz Silva, Valdilea G. Veloso, Paula M. Luz, and Beatriz Grinsztejn. 2021. “Evidence of an Untamed HIV Epidemic among MSM and TGW in Rio de Janeiro, Brazil: A 2018 to 2020 Cross-Sectional Study Using Recent Infection Testing.” Journal of the International AIDS Society 24 (6): e25743.

Vandormael, Alain, Adrian Dobra, Till Bärnighausen, Tulio de Oliveira, and Frank Tanser. 2018. “Incidence Rate Estimation, Periodic Testing and the Limitations of the Mid-Point Imputation Approach.” International Journal of Epidemiology 47 (1): 236–45.

Voetsch, Andrew C., Yen T. Duong, Paul Stupp, Suzue Saito, Stephen McCracken, Trudy Dobbs, Frieda S. Winterhalter, et al. 2021. “HIV-1 Recent Infection Testing Algorithm With Antiretroviral Drug Detection to Improve Accuracy of Incidence Estimates.” Journal of Acquired Immune Deficiency Syndromes 87 (Suppl 1): S73–80.

Wawer, M. J., N. K. Sewankambo, D. Serwadda, T. C. Quinn, L. A. Paxton, N. Kiwanuka, F. Wabwire-Mangen, et al. 1999. “Control of Sexually Transmitted Diseases for AIDS Prevention in Uganda: A Randomised Community Trial. Rakai Project Study Group.” The Lancet 353 (9152): 525–35.

Wood, Derrick E., and Steven L. Salzberg. 2014. “Kraken: Ultrafast Metagenomic Sequence Classification Using Exact Alignments.” Genome Biology 15 (3): R46.

Wu, Julia Wei, Oscar Patterson-Lomba, Vladimir Novitsky, and Marcello Pagano. 2015. “A Generalized Entropy Measure of Within-Host Viral Diversity for Identifying Recent HIV-1 Infections.” Medicine 94 (42). https://doi.org/10.1097/MD.0000000000001865.

Wymant, Chris, François Blanquart, Tanya Golubchik, Astrid Gall, Margreet Bakker, Daniela Bezemer, Nicholas J. Croucher, et al. 2018. “Easy and Accurate Reconstruction of Whole HIV Genomes from Short-Read Sequence Data with Shiver.” Virus Evolution 4 (1): vey007.

Wymant, Chris, Matthew Hall, Oliver Ratmann, David Bonsall, Tanya Golubchik, Mariateresa de Cesare, Astrid Gall, Marion Cornelissen, Christophe Fraser, and STOP-HCV Consortium, The Maela Pneumococcal Collaboration, and The BEEHIVE Collaboration. 2018. “PHYLOSCANNER: Inferring Transmission from Within- and Between-Host Pathogen Genetic Diversity.” Molecular Biology and Evolution 35 (3): 719–33.

Zanini, Fabio, Johanna Brodin, Lina Thebo, Christa Lanz, Göran Bratt, Jan Albert, and Richard A. Neher. 2015. “Population Genomics of Intrapatient HIV-1 Evolution.” eLife 4 (December). https://doi.org/10.7554/eLife.11282.

Zheng, Qi, Susan Ruone, William M. Switzer, Walid Heneine, and J. Gerardo García-Lerma. 2012. “Limited SHIV Env Diversification in Macaques Failing Oral Antiretroviral Pre-Exposure Prophylaxis.” Retrovirology 9 (May): 40.

Zhou, Shuntai, Sabrina Sizemore, Matt Moeser, Scott Zimmerman, Erika Samoff, Victoria Mobley, Simon Frost, et al. 2021. “Near Real-Time Identification of Recent Human Immunodeficiency Virus Transmissions, Transmitted Drug Resistance Mutations, and Transmission Networks by Multiplexed Primer ID-Next-Generation Sequencing in North Carolina.” The Journal of Infectious Diseases 223 (5): 876–84.

